# The impact of the COVID-19 pandemic on health service utilisation following self-harm: a systematic review

**DOI:** 10.1101/2022.01.26.22269901

**Authors:** Sarah Steeg, Ann John, David Gunnell, Nav Kapur, Dana Dekel, Lena Schmidt, Duleeka Knipe, Ella Arensman, Keith Hawton, Julian PT Higgins, Emily Eyles, Catherine Macleod-Hall, Luke A McGuiness, Roger T Webb

**Author notes:** Contributed equally (Ann John, David Gunnell).

## Abstract

**Background:** Evidence on the impacts of the pandemic on healthcare presentations for self-harm has accumulated rapidly. However, existing reviews do not include studies published beyond 2020.

**Aims:** To systematically review evidence on health services utilisation for self-harm during the COVID-19 pandemic.

**Methods:** A comprehensive search of multiple databases (WHO COVID-19 database; Medline; medRxiv; Scopus; PsyRxiv; SocArXiv; bioRxiv; COVID-19 Open Research Dataset, PubMed) was conducted. Studies reporting presentation frequencies for self-harm published from 1^st^ Jan. 2020 to 7^th^ Sept. 2021 were included. Study quality was assessed using a critical appraisal tool.

**Results:** Fifty-one studies were included. 59% (30/51) were rated as ‘low’ quality, 29% (15/51) as ‘moderate’ and 12% (6/51) as ‘high-moderate’. Most evidence (84%, 43/51 studies) was from high-income countries. 47% (24/51) of studies reported reductions in presentation frequency, including all 6 rated as high-moderate quality, which reported reductions of 17- 56%. Settings treating higher lethality self-harm were overrepresented among studies reporting increased demand. Two of the 3 higher quality studies including study observation months from 2021 reported reductions in service utilisation. Evidence from 2021 suggested increased use of health services following self-harm among adolescents, particularly girls.

**Conclusions:** Sustained reductions in service utilisation were seen into the first half of 2021. However, evidence from low- and middle-income countries is lacking. The increased use of health services among adolescents, particularly girls, into 2021 is of concern. Our findings may reflect changes in thresholds for help seeking, use of alternative sources of support and variable effects of the pandemic across different groups.

## Introduction

The COVID-19 pandemic has led to deterioration in population mental health and has placed considerable additional strains on health systems. ^1^ ^2^ The pandemic has also heightened many of the risk factors for suicidal behaviour, such as job insecurity and unemployment, access to food, education and healthcare and the availability of family and community support. ^3^ Understanding and quantifying trends in help seeking for self-harm is a vital part of the public mental health response to COVID-19. It could help expound the apparent paradox observed during the early stages of the pandemic; while population mental health deteriorated, ^4^ fewer people sought help for their mental health from primary and secondary care services. ^5^ Examining self-harm presentations across health settings could help understand longer-term population impacts and inform planning of services and interventions in the future phases of the pandemic.

Numerous studies from high-income countries reported marked reductions in health service utilisation during the second quarter of 2020 following the start of the COVID-19 pandemic. For example, considerable reductions in diagnoses for acute physical and mental illnesses were found in the UK following introduction of the national lockdown in March 2020, with only partial recovery by July 2020. ^6^ In another UK study, reductions of around a third in health service contacts specifically for self-harm were found. ^7^ Focussing specifically on hospital admission for self-harm, overall reductions of just over 8% were reported in France, though increases in more serious potentially lethal acts of self-harm were observed. ^8^ Evidence relating to the indirect health impacts resulting from the pandemic in low- and middle-income countries also suggests care for non-communicable diseases and mental disorders has been severely disrupted. ^9^ A systematic review on the impact of the pandemic on suicide and self-harm in low- and middle-income countries found mixed evidence, with either a decrease or no discernible impact in reported self-harm episodes along with increases in certain age groups. ^10^

In 2020, a living systematic review was established to provide an up-to-date resource and data synthesis of evidence on the impact of the COVID-19 pandemic on self-harm and suicidal behaviour. ^11^ The most recent update of the review included studies up to 19^th^ October 2020 and included 20 health service utilisation studies, including 11 focussing specifically on health service contact following self-harm/suicide attempts. ^12^ The review reported that most studies reported a decrease in presentations to health services for self-harm during the early months of the COVID-19 pandemic.

However, all 20 studies were of high-income countries and the latest month of observation was August 2020. ^13–15^ In the subsequent months many health services adapted and ‘stay at home’ orders eased, although these restrictions later returned in many countries and regions. While studies suggest service utilisation had returned to expected volumes in some countries by the third quarter of 2020, ^12^ it is not known how subsequent restrictions and ongoing pressures on health systems in response to further waves of COVID-19 affected help-seeking and access to healthcare for self-harm. In this article we report on evidence concerning the frequency (reported incident or prevalent episode counts or rates) of health service utilisation for self-harm after the onset of the pandemic compared to before the pandemic. There has been no synthesis of studies published since October 2020, some of which would be expected to include the later observation periods covering the latter months of 2020 and first half of 2021, as the pandemic continued to affect populations globally. Our aim was to systematically identify, review and synthesise evidence relating to utilisation of health services for self-harm since the COVID-19 pandemic began in the first quarter of 2020.

## Methods

The protocol for the methodology applied in conducting the systematic review is registered within a living systematic review of the impact of the COVID-19 pandemic on self-harm and suicidal behaviour (PROSPERO ID CRD42020183326; registered on 1st May 2020). ^11^ ^5^ ^12^ Additional inclusion and exclusion criteria specific to our research question were applied and further screening, data extraction and study quality assessments were conducted. To address our research question, ‘did the frequency of health service presentation for self-harm during the pandemic change compared to antecedent periods?’, we applied the following inclusion and exclusion criteria:

### Inclusion criteria

- Published from 1^st^ Jan. 2020 to 7^th^ Sept. 2021.
- Written in any language.
- Investigation of health service utilisation among the general population
  - Including presentations to general hospital emergency departments (EDs), primary healthcare services, specialist mental healthcare services (accessible to general population), other secondary healthcare services that treat people who have self-harmed / attempted suicide (e.g. surgery) and admission to hospitals.
- Outcomes were presentations for self-harm, including:
  - broad definition of self-harm (defined as non-fatal intentional self-injury, intentional self-poisoning involving drugs or non-ingestible substances, including non-suicidal acts) or attempted suicide, including hospital attendance and/or admission for these reasons ^11^
  - narrower definition, for example studies focussed only on suicide attempts or specific methods of self-harm.
- Comparisons:
  - health service presentation frequencies (including incident or prevalent episode counts or rates) for self-harm before and after the beginning of the COVID-19 pandemic, considering specific time periods separately; for example, both initial and subsequent lockdown periods.

### Exclusion criteria

- Studies without pre-pandemic observation periods or measurements, including those reporting use of service initiatives implemented in response to the pandemic, with no pre-pandemic comparison period.
- Reports where only an abstract was available.
- Studies focussing on specific groups such as those with a specific physical of psychiatric diagnosis (including COVID-19) or where the baseline population was existing patients within a specialist service, such as psychiatric inpatients.
- Studies reporting self-harm and suicidal thoughts as a combined measure.
- Studies reporting proportions of self-harm presentations, without reporting absolute figures
- Studies of suicides.

### Data analysis

The list of studies used for screening was obtained from the main living systematic review database. ^12^ This database is updated automatically using daily electronic searches of multiple databases (WHO COVID-19 database; Medline; medRxiv; Scopus; PsyRxiv; SocArXiv; bioRxiv; COVID-19 Open Research Dataset, PubMed) (see Supplement 2 for search strategy for each database). Screening was conducted in two stages: the citations returned by the automated searches were assessed by four screeners (CO, EE, DD, CM-H) to identify potentially relevant studies, then AJ, DG, DK or RW assessed the full text of the studies to identify studies to be included in the main living systematic review. In addition, expert reviewers (AJ, DG, DK and RW) completed daily assessments of the automated results, which included basic data extraction and assigning studies manually to a study design category, along with a description of the study design.

Identification and screening of studies for the current review was conducted using a methodology developed as part of an existing living systematic review (Fig. 1). ^12^ Studies included publications identified in the living systematic review from 1^st^ Jan. 2020 up to 7^th^ Sept. 2021. Screening was conducted according to the inclusion and exclusion criteria for the current review. The list of studies was extracted from the main living systematic review database on 14^th^ Sept. 2021. Categories assessed for inclusion in the current review were ‘service utilisation’, ‘before/after studies’, ‘time trends analysis’ and ‘examination of electronic health records’ (Fig. 1). ^16^

**Figure 1:**
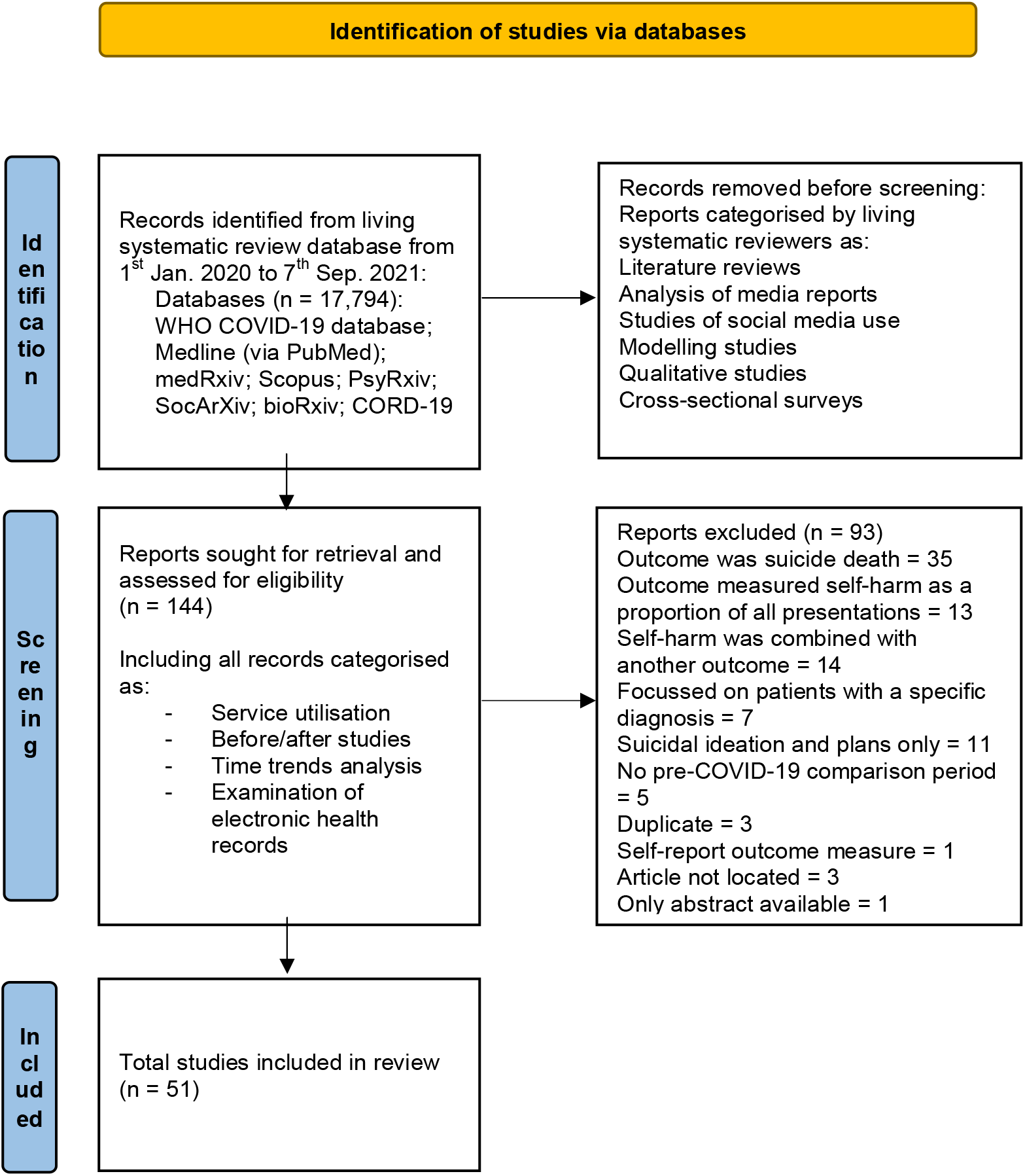
PRISMA flow diagram ^12^

As part of the identification and screening procedure, further screening and data extraction was completed for the current systematic review using a proforma designed to collect standardised information from each study (Table S1). Study quality and risks of bias were assessed using an adapted version of an existing National Institute for Health (NIH) quality assessment tool, designed specifically for studies using before and after designs. ^17^ The NIH tool was adapted by authors DK, JH and DG to include consideration of the pandemic and associated lockdown periods and other societal restrictions as the intervention of interest and to account for the use of health service data sources in the study designs. The overall assessment tool was used to judge the quality of studies, with predefined criteria established for studies to be rated as high or moderate quality. Screening, data extraction and quality assessments were conducted by SS. A second rater (DD) assessed eligibility for 20% of the studies sought for retrieval and conducted independent data extraction and analysis on 10% of the included studies. There was agreement on all eligibility assessments and study quality ratings. If a source was not available in English, data extraction was conducted by expert reviewers fluent in the language that the article was written in. Where included studies were preprints, searches for peer-reviewed version were conducted and the updated peer-reviewed version was used for data extraction where available. Data synthesis was conducted by extracting, assessing and tabulating key aspects of the studies, including setting, study design, data sources, outcome measures, follow-up and comparison periods, main findings and study quality. The main effect measure of interest was percentage difference in presentation frequency during a defined COVID-19 period compared to a pre-COVID-19 comparison period. If this data were missing, the overall direction of change (e.g. increase/no change/decrease) was recorded. Higher quality studies were prioritised and reported separately during data synthesis and presentation of results.

## Results

### Description of included studies

Fifty-one studies were included. These were from healthcare settings including general hospital EDs (39%, 20/51), trauma and surgery admissions (22%, 11/51), children’s hospitals (8%, 4/51), primary care (8%, 4/51), general hospital admissions (6%, 3/51), paediatric EDs (6%, 3/51), ambulance calls (4%, 2/51), liaison psychiatry referrals (4%, 2/51) paediatric trauma admissions (2%, 1/51) and a multiservice setting (2%, 1/51) (Table 1 and Table S1). Study quality was mixed; 59% (30/51) were rated as ‘low’ quality, 29% (15/51) as ‘moderate’ and 12% (6/51) as ‘high-moderate’. Reasons for studies being rated as low quality commonly included small event counts, absence of clearly defined patient eligibility criteria, and poorly described data extraction/collection methodology. Most of the evidence (84%, 43/51 studies) was from investigations conducted in high-income countries (Table 1). Forty-two of the 51 studies were reported in peer-reviewed articles, four were preprints, four were letters or editorials and one was a report.

**Table 1:**
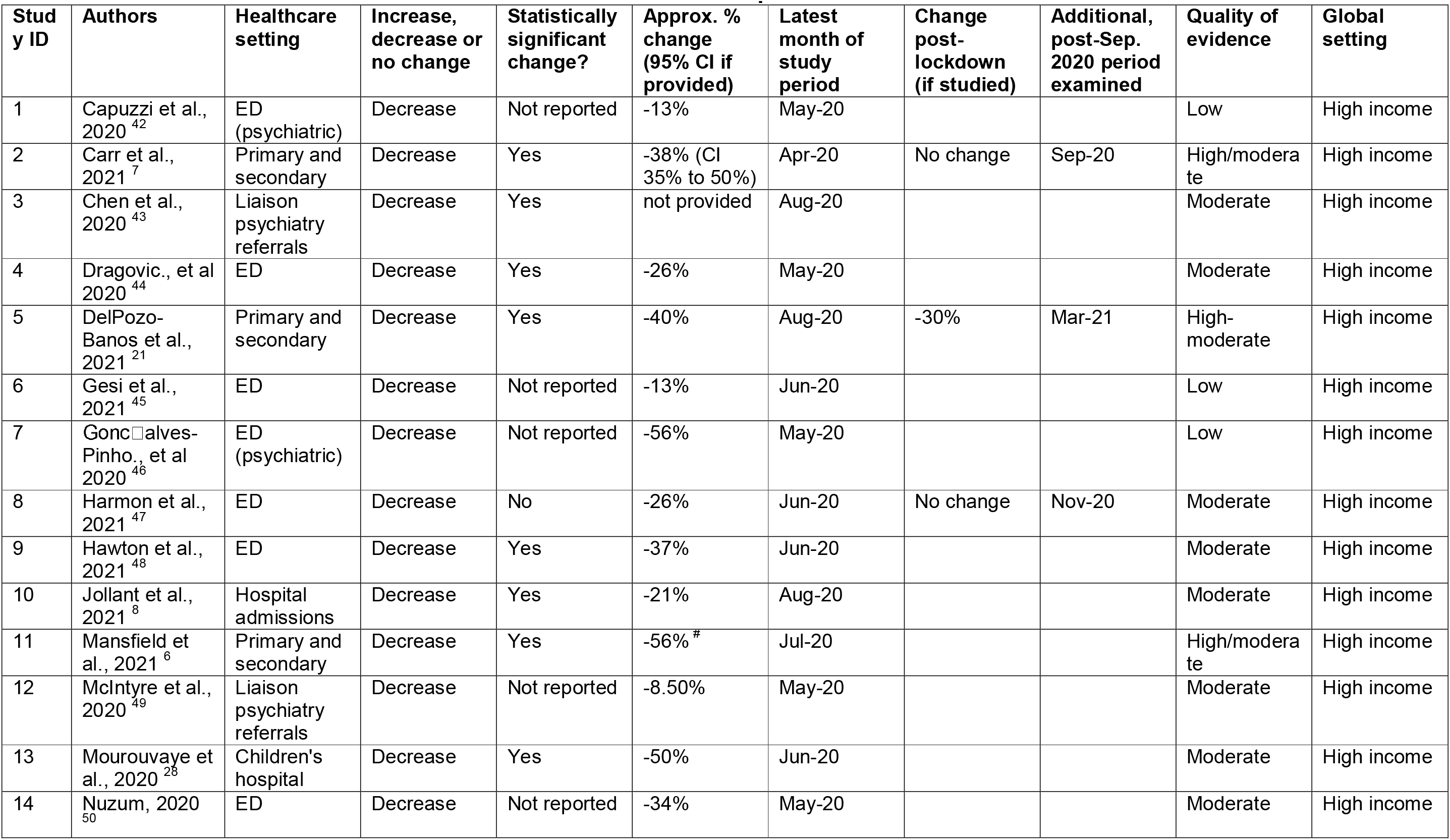
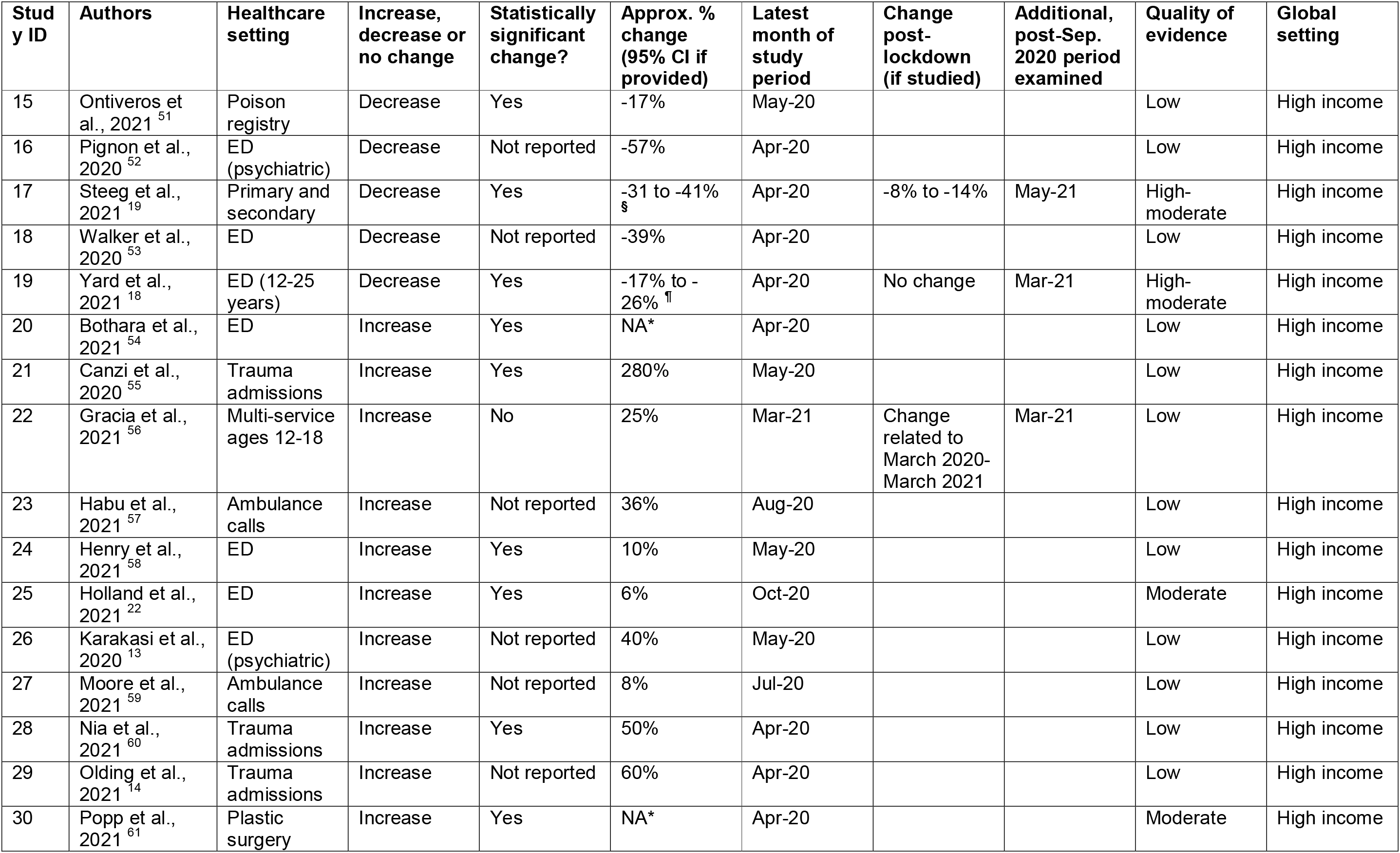
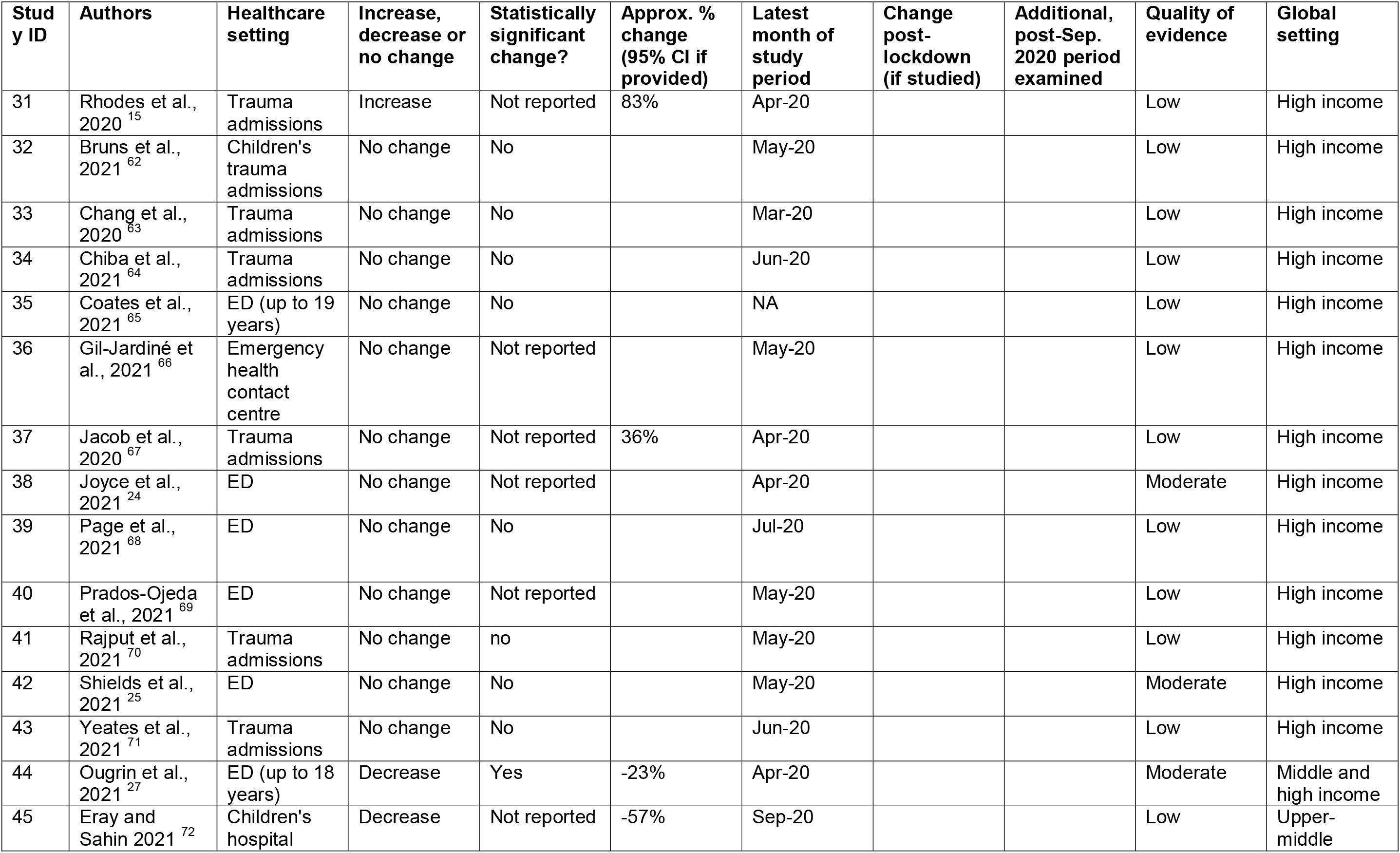
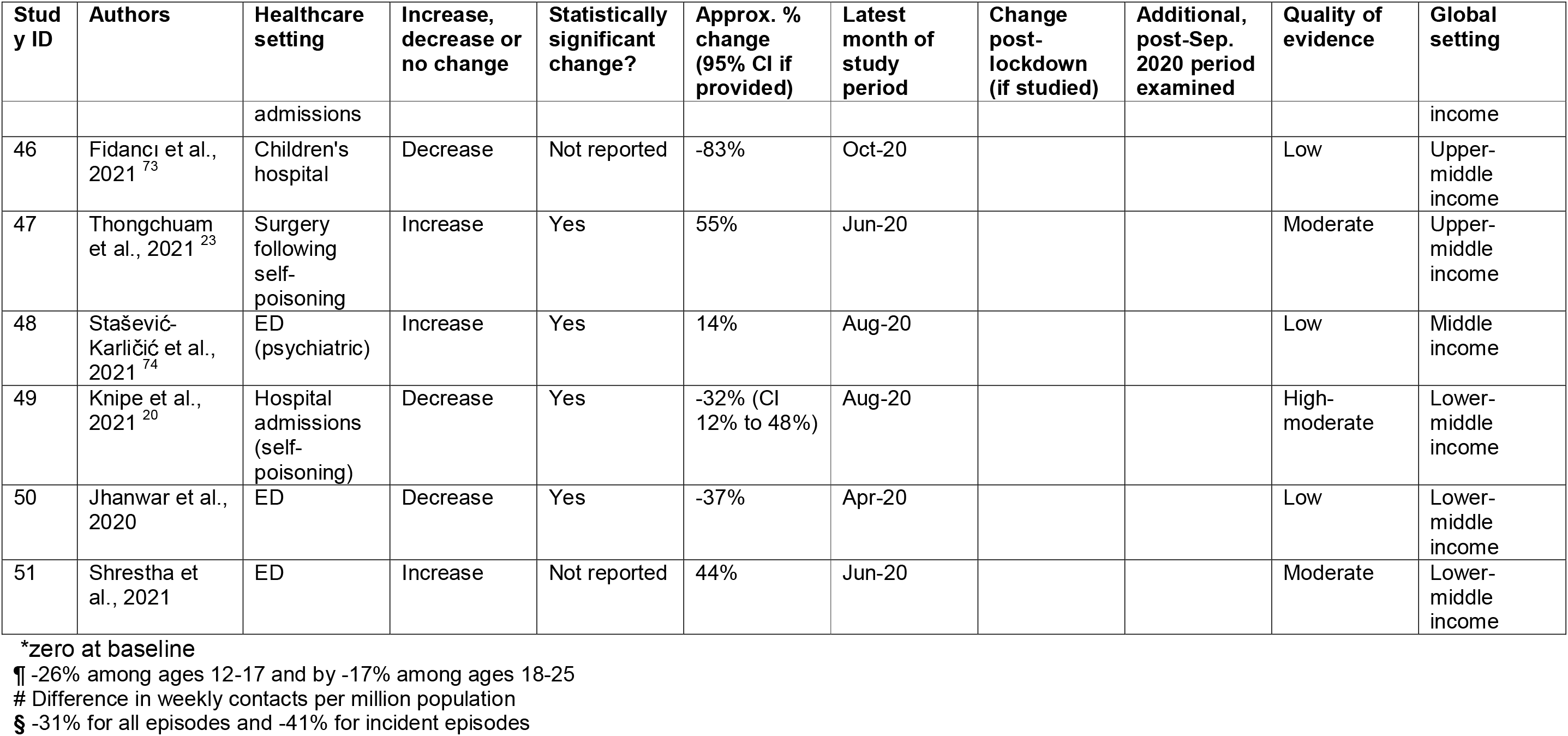
Characteristics of included studies: 1^st^ Jan. 2020 to 7^th^ Sep. 2021

### Findings of included studies

Almost half (47%, 24/51) of the studies reported reductions in presentation frequency (Figure 2) for the duration of the period studied, the majority of which included months no later than August 2020. All 6 studies rated as high-moderate quality found decreases in service utilisation during the early months of the pandemic, with reductions of between 17 and 56% reported. ^6^ ^7^ ^18–21^ These studies were of primary and secondary care settings combined (4 studies), ED presentations among ages 18 to 25 years (1 study) and self-poisoning presentations to hospital (1 study). Four studies used healthcare records in the UK to compare expected vs. observed primary and secondary-care recorded episodes of self-harm, and found reductions of between 26 and 44%. ^6^ ^7^ ^19^ ^21^ Another study based in Sri Lanka found a 32% reduction in hospital presentations for self-poisoning compared to pre-pandemic numbers. However, these estimates included months no later than August 2020.

**Figure 2:**
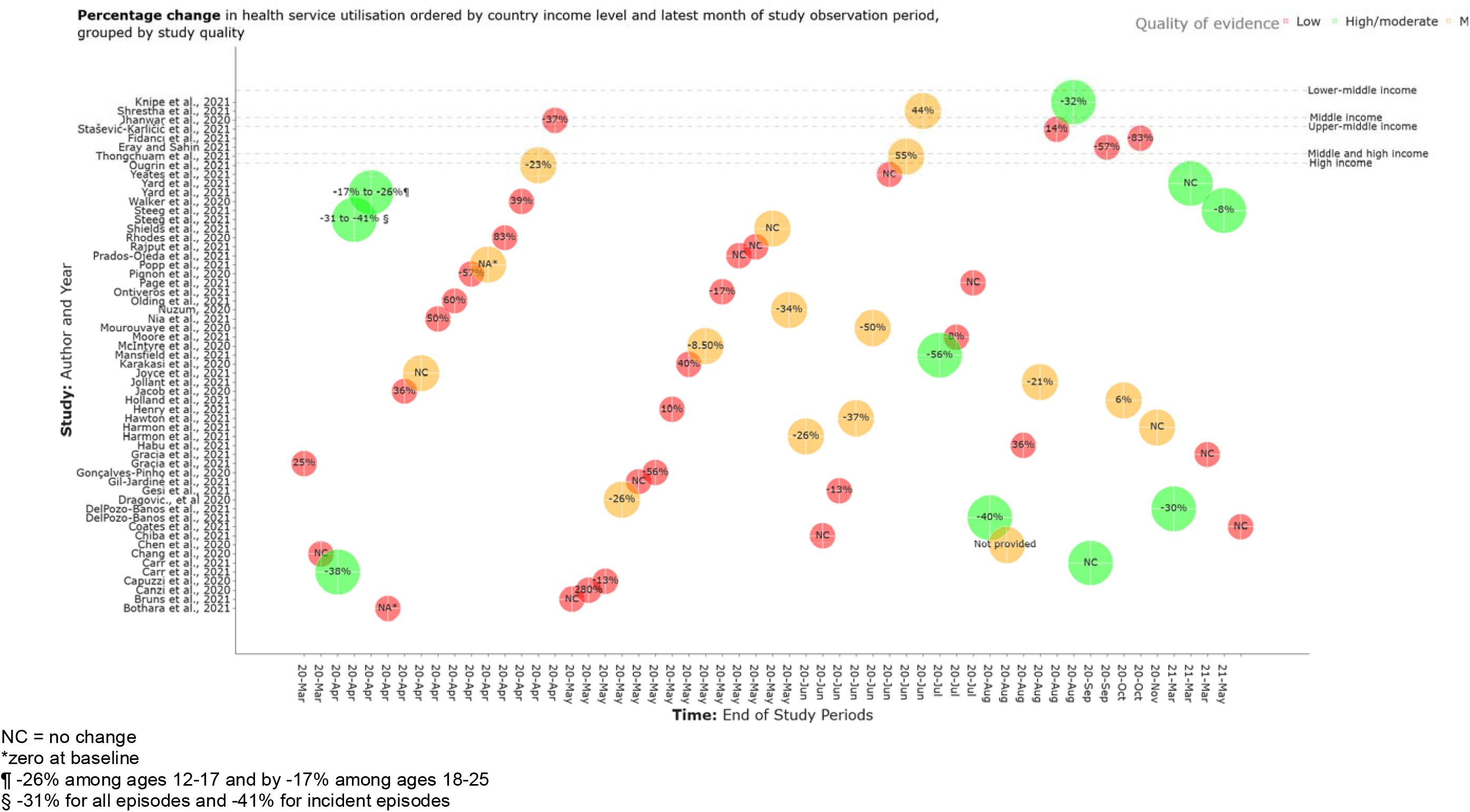
Percentage change in health service utilisation ordered by country income level and latest month of study observation period, grouped by study quality.

Five studies used national or nationally representative data. Four of these were assessed as high-moderate quality and reported decreases in service utilisation of between 26 and 56%. One moderate quality study reported a 6% increase in ED presentations. ^22^ This US-based study only included self-harm episodes classified as suicide attempts, therefore may not reflect service use for self-harm more broadly.

Increases were reported in 15/51 (29%) studies, none of which were assessed as being of high-moderate quality and 4 were rated as moderate quality. An examination of the number of people admitted to a surgical department following self-harm by ingestion of corrosive substances was found to increase by 55% in one Bangkok hospital, though numbers in the study were relatively low. ^23^ Other moderate quality studies reporting increased patient numbers included ED and surgery services, settings likely to be encountering patients with more medically severe episodes of self-harm.

Twelve out of 51 (24%) studies reported no change in service utilisation, including no high-moderate quality studies and 2 assessed as moderate quality. These were both conducted in ED settings, with one New Zealand ED reporting no change in self-harm presentations ^24^ and a UK-based study reporting no change in hospital admission following ED presentations for self-harm ^25^ A further 6 studies were conducted in trauma settings, though all were rated as low quality.

Most studies (46/51) included up to a maximum of 8 months of follow-up from the first wave of the pandemic (March to October 2020). Among the 4 studies including months from 2021 in their observation period (up to May 2021), 3 were rated as high-moderate quality. Among these, 2 studies of primary and secondary care-recorded self-harm reported longer-term reductions of between 8 and 30% respectively ^19^ ^21^ and another study of ED presentations by young people aged 12 to 25 years found no overall change. ^18^ Studies including follow-up months beyond 2020 were limited to those originating from high-income countries.

### Findings by study settings and subgroups

7/51 (14%) studies were conducted in upper-middle-income (3 studies), middle-income (1) and lower-middle income (3) countries, one of which was rated as high-moderate quality. Four studies found a decrease in service use and three reported an increase. The study rated as high-moderate quality reported on self-poisoning episodes in a lower-middle-income setting; using health record data from a toxicology unit in a Sri Lankan hospital, a 32% reduction in hospital presentations for self-poisoning was found compared to pre-pandemic numbers. ^20^ A study of moderate quality conducted in one Nepalese ED found an increase of 44% in presentations for self-harm during the lockdown period compared to the same period the previous year, with indications that severity of self-harm was higher, though the numbers of presentations in both the lockdown and comparison periods were relatively small. ^26^

Eighteen studies included examination of service use for self-harm specifically among children and/or young people, with five rated as high-moderate quality. One high-moderate quality study including approximately 71% of the US’s EDs in 49 states examined presentations among ages 18 to 25 years and found reductions of 26% among ages 12-17 and 17% among ages 18-25 in April 2020. However, when examining presentation rates over the longer-term, through to March 2021, increases compared to equivalent weeks in 2019 were found for girls aged 12-17. Among boys aged 12-17 and all adults aged 18-25 years, rates through to March 2021 were in line with those in 2019. ^18^ Another high-moderate quality study, based in the UK, reported increased numbers of presentations to primary and secondary care among all adolescents aged 10-17 years up to May 2021. ^19^ These findings are in contrast to those reported in other moderate quality studies using earlier COVID-19 observation periods (up to June 2020) where younger people were found to have significantly fewer self-harm presentations than in the equivalent period in 2019. ^27^ ^28^

## Discussion

### Main findings

All of the studies assessed as high-moderate quality reported decreases in service utilisation following self-harm and were conducted in settings reflecting a broad spectrum of self-harm with higher frequency of presentations such as primary care. We found that settings treating episodes of self-harm with lower frequency and higher lethality, such as trauma admissions and ambulance calls, were overrepresented among studies that reported increased or no change in demand. Among higher quality studies that included months from 2021 in their observation period, numbers of people seeking help from health services were found to be either closer to pre-pandemic levels, though still lower than expected, or in line with expected numbers. Evidence from 2021 also suggested there was increased utilisation of health services following self-harm among adolescents, girls particularly so. However, studies including follow-up months from 2021 were limited to those originating from high-income countries.

### Strengths and limitations

This systematic review is the first to examine up-to-date evidence regarding associations between the COVID-19 pandemic and frequency of health service utilisation for self-harm. An established, peer-reviewed living systematic review methodology, ^11^ with ongoing data extraction by a panel of suicide prevention experts, was used as the basis for this review. This approach, along with a specific focus on studies comparing frequency in utilisation of health services following self-harm in different settings during the COVID-19 pandemic versus antecedent pre-pandemic periods, enables timely synthesis of the evolving evidence base.

The findings of our study should be interpreted with some important caveats in mind. We excluded 6 studies that reported self-harm and suicidal thoughts as a combined measure as it was not possible to make a like-for-like comparison with findings pertaining specifically to acts of self-harm. However, we included studies using a broad range of definitions of self-harm, including those that measured and reported on suicide attempts or self-poisoning methods only. We also did not include temporal trends in the proportion of all presentations that were for self-harm as a primary outcome, due to the limitation that this outcome would be affected by changes in the overall number of presentations for reasons other than self-harm.

We conducted a comprehensive narrative synthesis of the data rather than a meta-analysis due to heterogeneity in the pandemic and antecedent comparison periods, definitions of self-harm applied, and healthcare settings that studies were conducted in. Performing a meta-analysis will be considered for future updates of the living systematic review. The studies included in our review are of mixed quality and are greatly under-representative of middle- and low-income countries. While we have reported findings according to these characteristics, overall findings should be interpreted in light of these considerations.

### Implications and comparison with existing evidence

Most studies came from high-income countries. Findings from higher quality studies suggested either there were continued reductions in health service utilisation into 2021, though to a lesser extent than earlier months of the pandemic, or that service use had broadly returned to pre-pandemic levels. However, these findings cannot necessarily be generalised to low- and middle-income countries. For example, allocation of COVID-19 vaccinations has been disproportionately skewed towards high-income countries. ^29^

Consequently, many low- and middle-income countries have experienced major subsequent waves of COVID-19 well into 2021. ^30^ The effects of these further waves of infection on many of the factors associated with self-harm - for example, unemployment, mental and physical ill health, poor access to healthcare - are likely to be considerable. ^31^ Subsequent waves of COVID-19 have also been experienced by high-income countries into the latter half of 2021. For example, from November 2021, some European countries introduced further societal restrictions. ^32^ Continued surveillance is therefore needed in all settings.

Our findings are consistent with reports of increased acuity of presentations in some mental health services. ^33^ ^34^ The increases in presentation frequency reported by studies that were conducted in healthcare settings treating more potentially lethal episodes of self-harm, such as ambulance calls and trauma admissions indicates that the pandemic has impacted the threshold for help seeking. Evidence also shows that non-statutory mental health services, such as charities, experienced increased demand in the months following the onset of the pandemic. ^35^ This may explain the apparent paradox observed during the first year of the pandemic, where deterioration in population mental health alongside reductions in health services utilisation was observed. This indicates that reductions seen in settings capturing a broader spectrum of self-harm do not simply reflect decreased incidence of self-harm or reduced clinical need. People who have harmed themselves non-fatally have a markedly elevated suicide risk subsequently, irrespective of self-harm method at the index episode, and degrees of suicidal intent can fluctuate between different self-harm episodes by the same person. ^36^ Therefore, it is vital that people harming themselves receive clinical intervention and that health services across the world work to ensure services are available to provide timely and accessible care. ^37^ ^38^

Studies examining changes in proportions of groups presenting with certain characteristics, and those examining combined ‘suicidal thoughts and self-harm’ outcomes, were not included in this systematic review as we were interested in absolute numbers of people using health services for self-harm. However, such studies can provide valuable information about help seeking behaviour in different groups. For example, a study of hospital attendance for suicidal ideation and self-harm in Australia’s Gold Coast region identified a number of groups with particularly reduced likelihood of presentation during March to August 2020, including Indigenous Australians and individuals with less severe suicidal and self-harm, while people younger than 18 years had increased numbers of presentations. ^39^

Another study conducted in a paediatric ED in New York City, USA found that while overall there were significant decreases in emergency attendances, visits for suicidal ideation and self-harm among young people increased. ^40^ Increases in numbers of adolescents referred to mental health services in Ireland were found from September 2020, following initial decline in April 2020. ^41^ Our findings of increased utilisation of health services for self-harm into the early months of 2021 among adolescents, particularly girls, within this context, are concerning and warrant urgent attention.

## Conclusions

All high-quality studies reported a fall in attendance frequency for self-harm during the early months of the pandemic. New evidence relating to the first and second quarters of 2021 indicated that longer-term impacts on health services were less marked than during the first wave of the pandemic, though reductions in frequency of presentation versus expected levels persisted. These patterns likely reflect changes in thresholds for help seeking, increases in frequency of higher acuity episodes of self-harm and increased use of non-statutory health services. The increased utilisation of health services among adolescents, particularly girls, into the early months of 2021 warrants particular attention. However, evidence from low- and middle-income countries is still limited. High-quality, multi-centre studies examining the longer-term impacts on health service utilisation for self-harm, particularly in low- and middle-income countries, including observation periods into 2021 and among children and young people, are urgently needed.

## Supporting information

PRISMA

## Data Availability

All data produced in the present study are available upon reasonable request to the authors.

## Declaration of Interest

DG, KH and NK are members of the Department of Health and Social Care (England) National Suicide Prevention Strategy Advisory Group.

## Funding

SS is funded by a University of Manchester Presidential Fellowship. NK and RTW are funded by the National Institute for Health Research (NIHR) Greater Manchester Patient Safety Translational Research centre.

## Acknowledgements

The authors thank Dr Claire Huish, Dr Florian Walter and Dr Laszlo Trefan for conducting data extraction for non-English language studies.

## Author contributions

Conceptualisation and design of study: SS, AJ, DG, RTW

Initial screening: EE, DD, CMH, DK, AJ, RTW, DG

Expert reviewing: AJ, DG, DK, RTW

Quality assessments: SS, DD

Data analysis: SS, DD, LS

Writing manuscript: SS

Critical reviewing and editing of manuscript: all authors

---

**Table S1:**
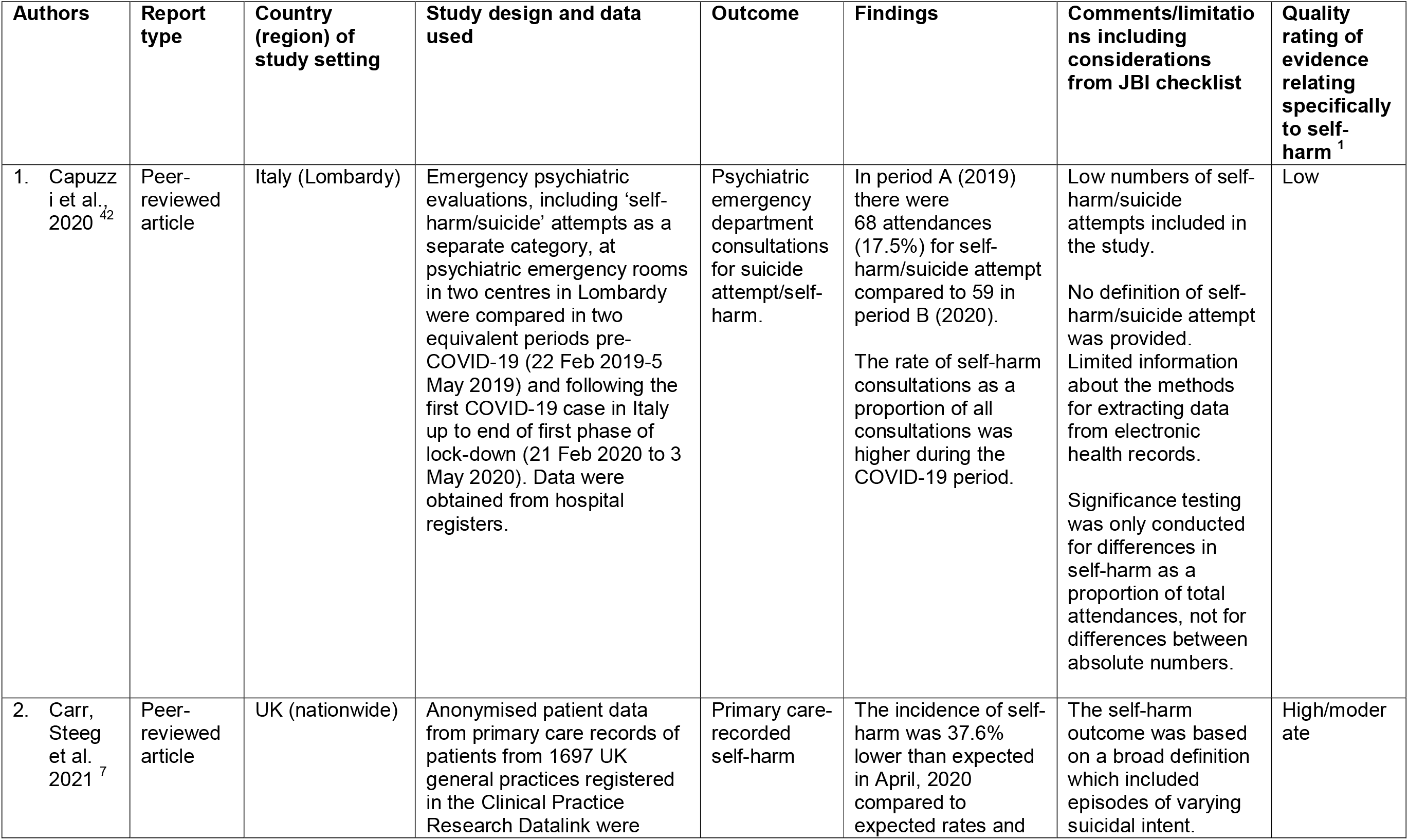
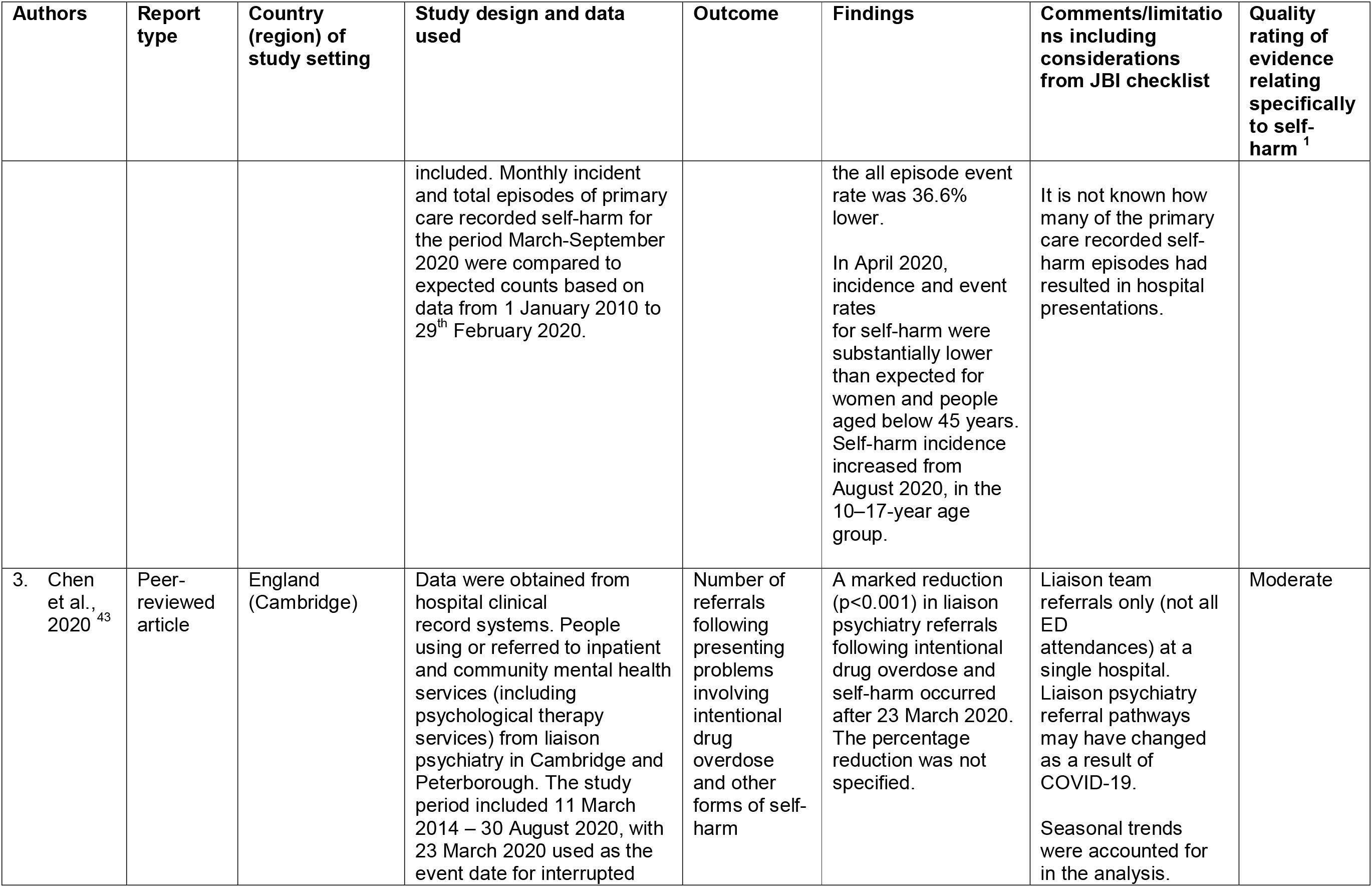
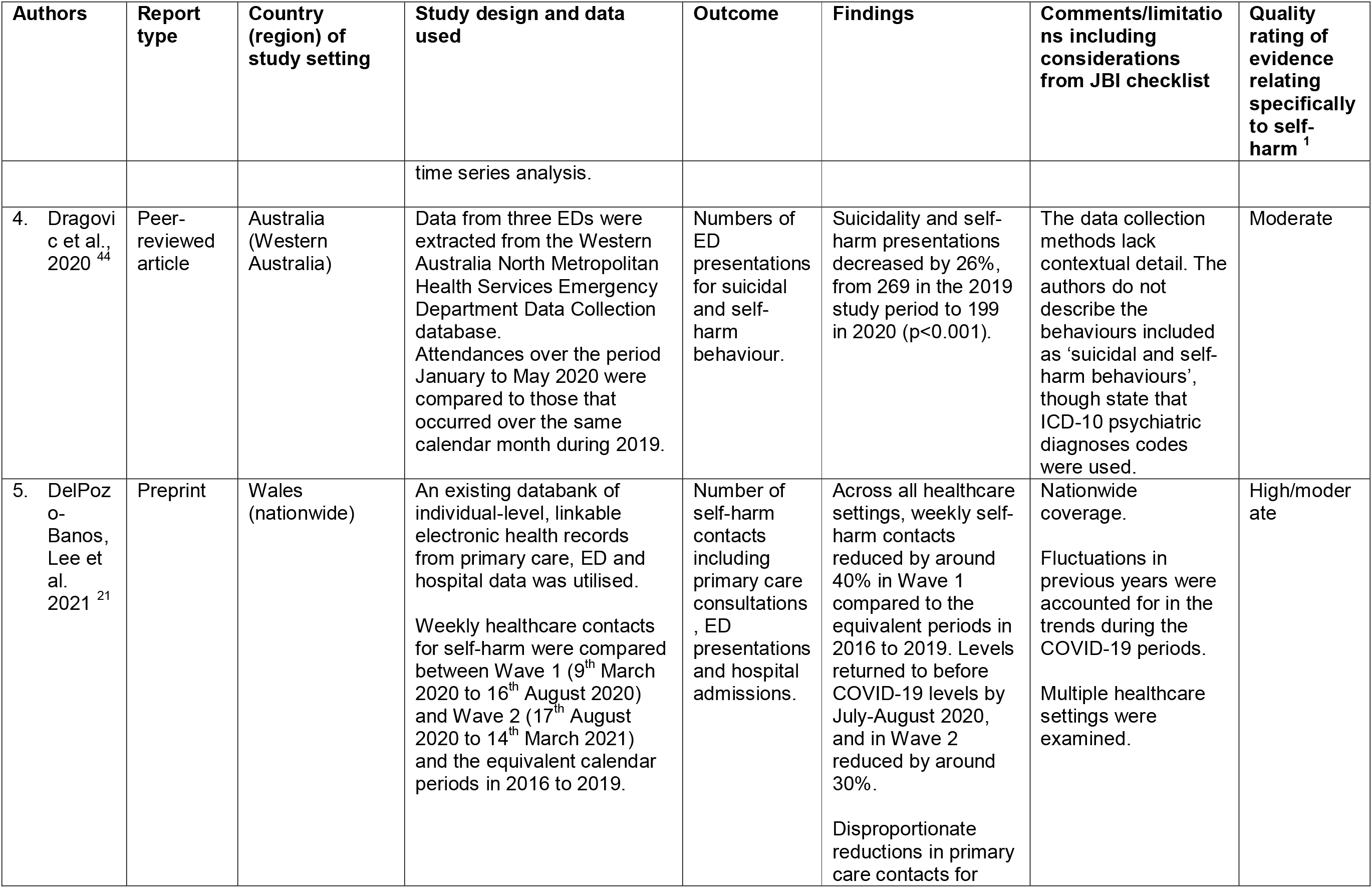
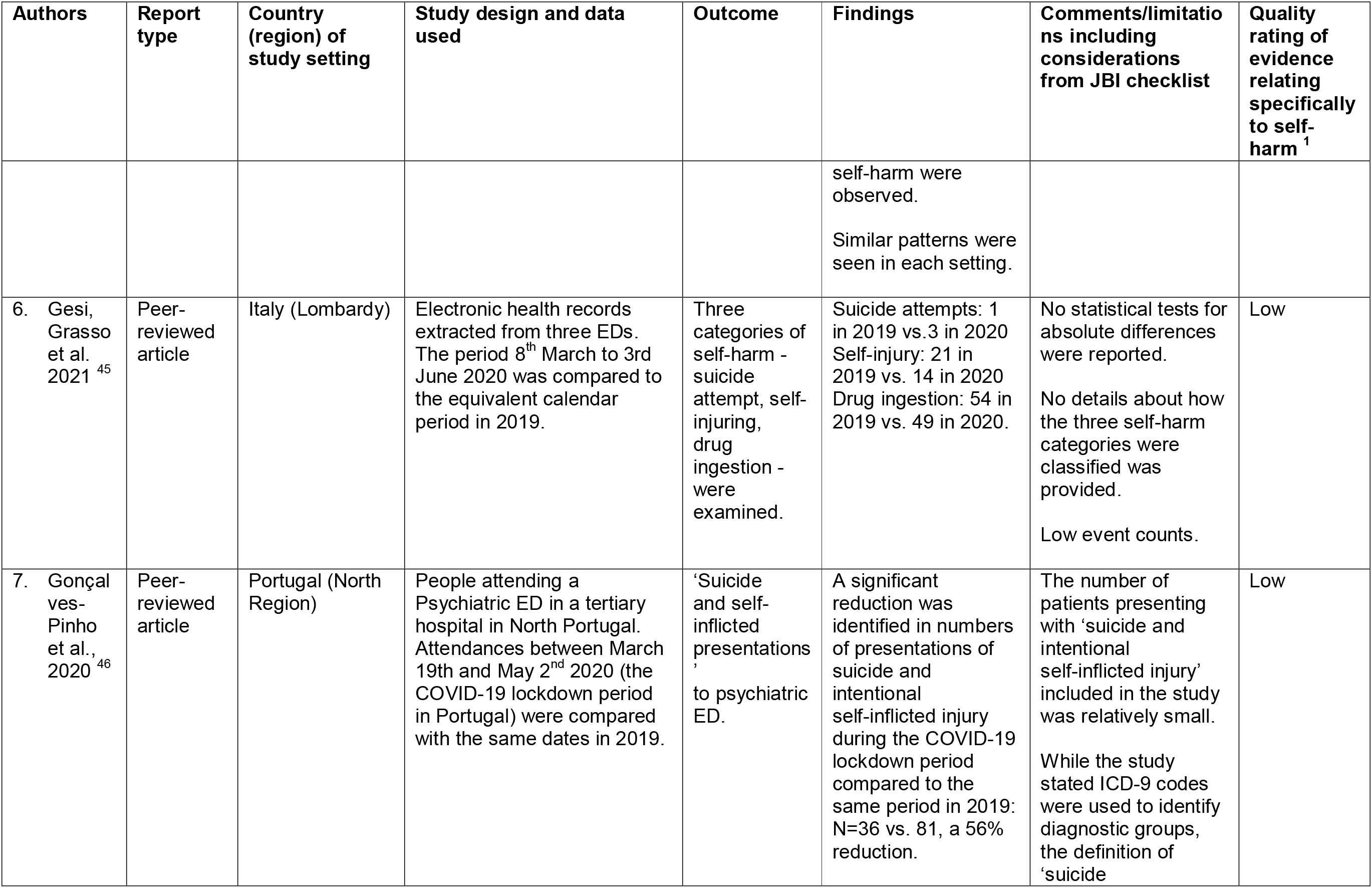
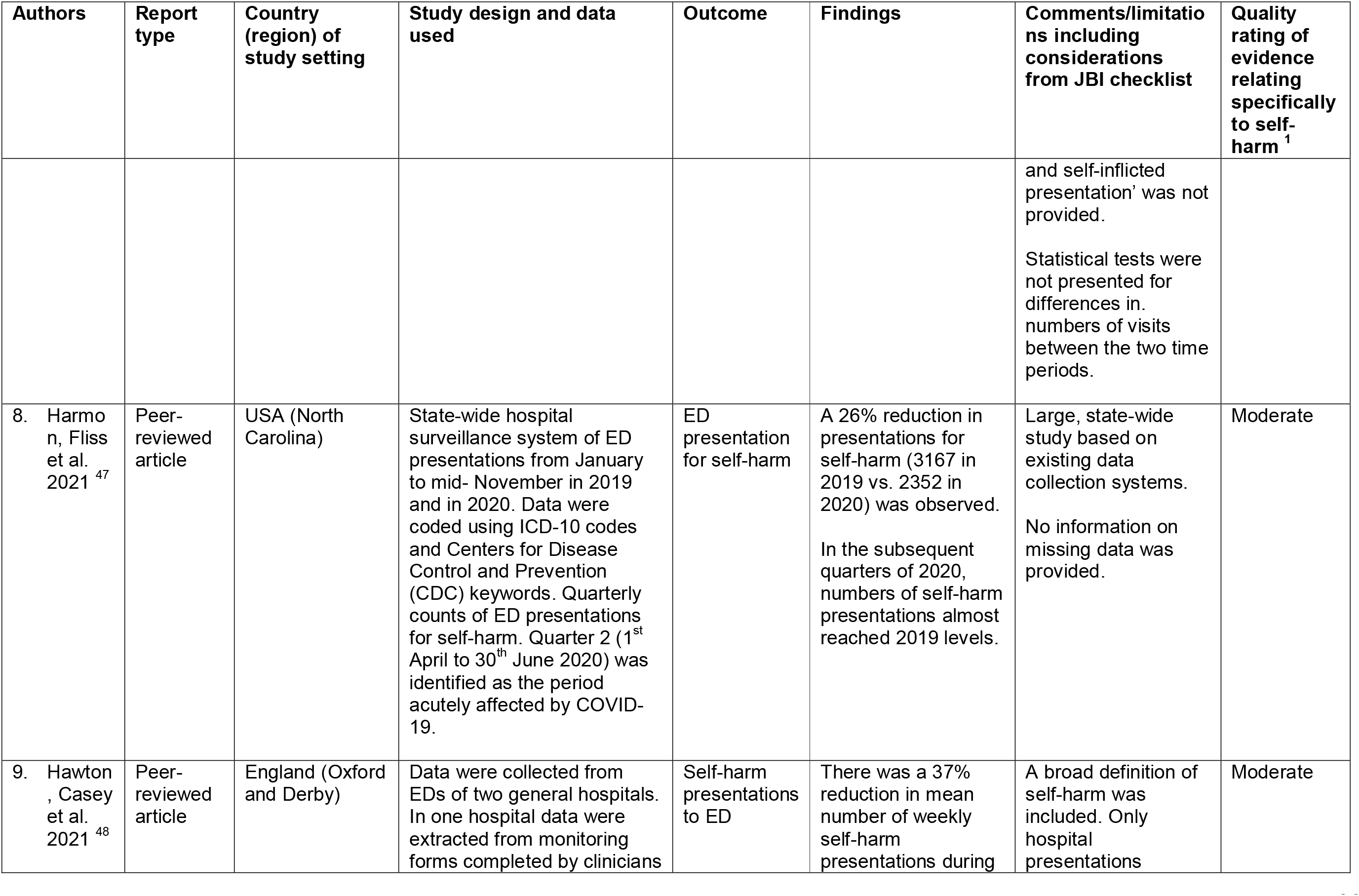
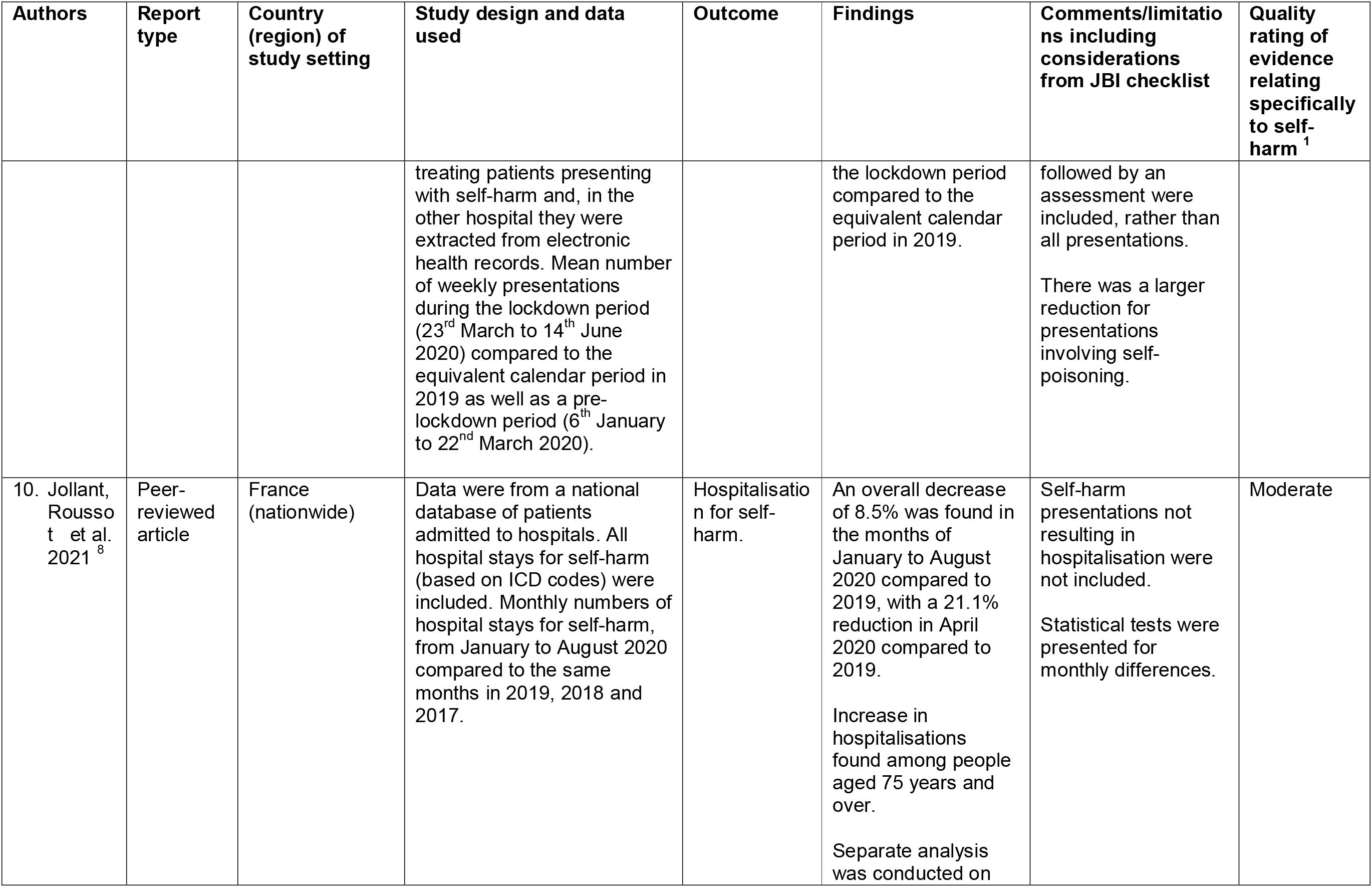
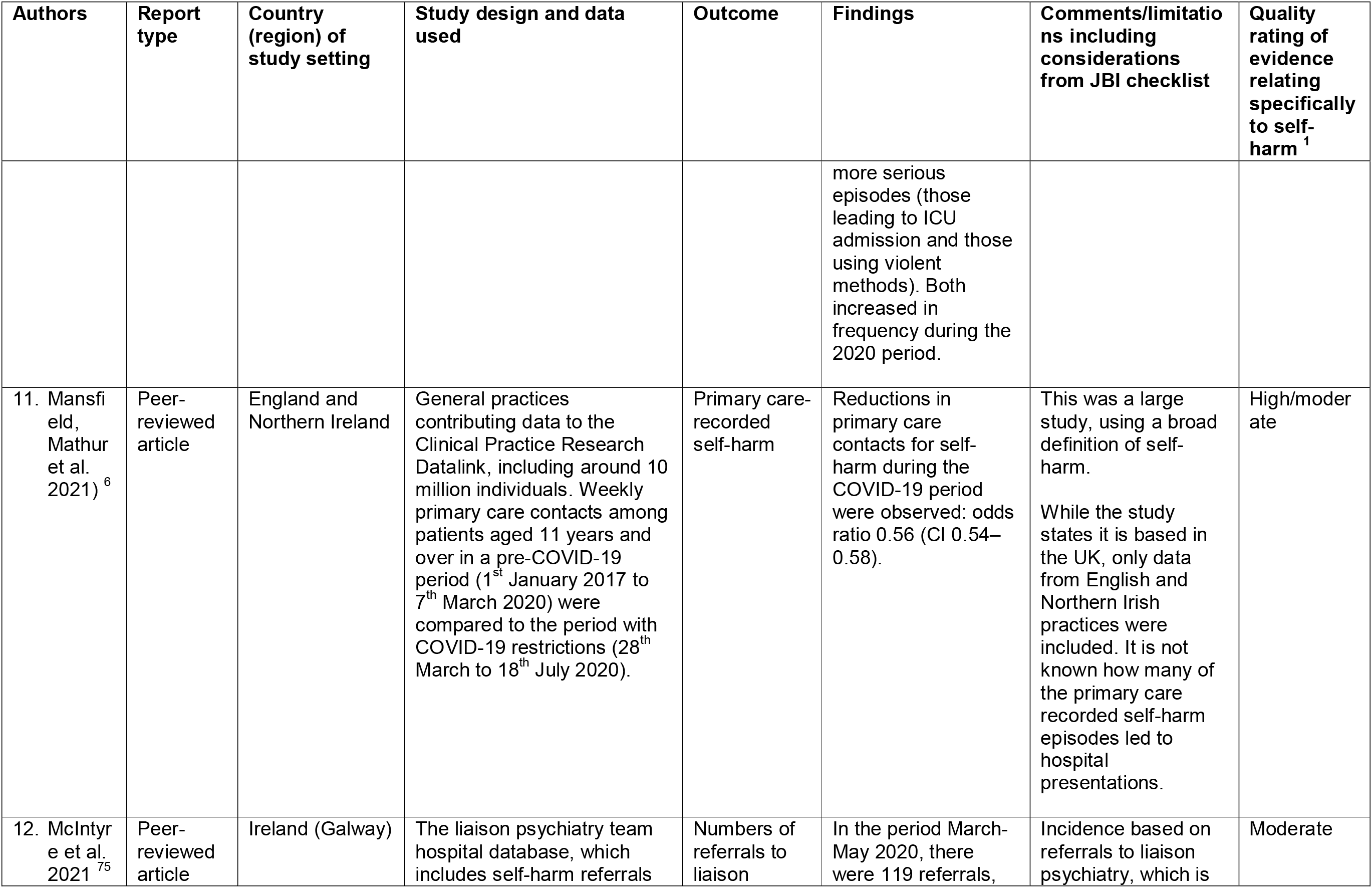
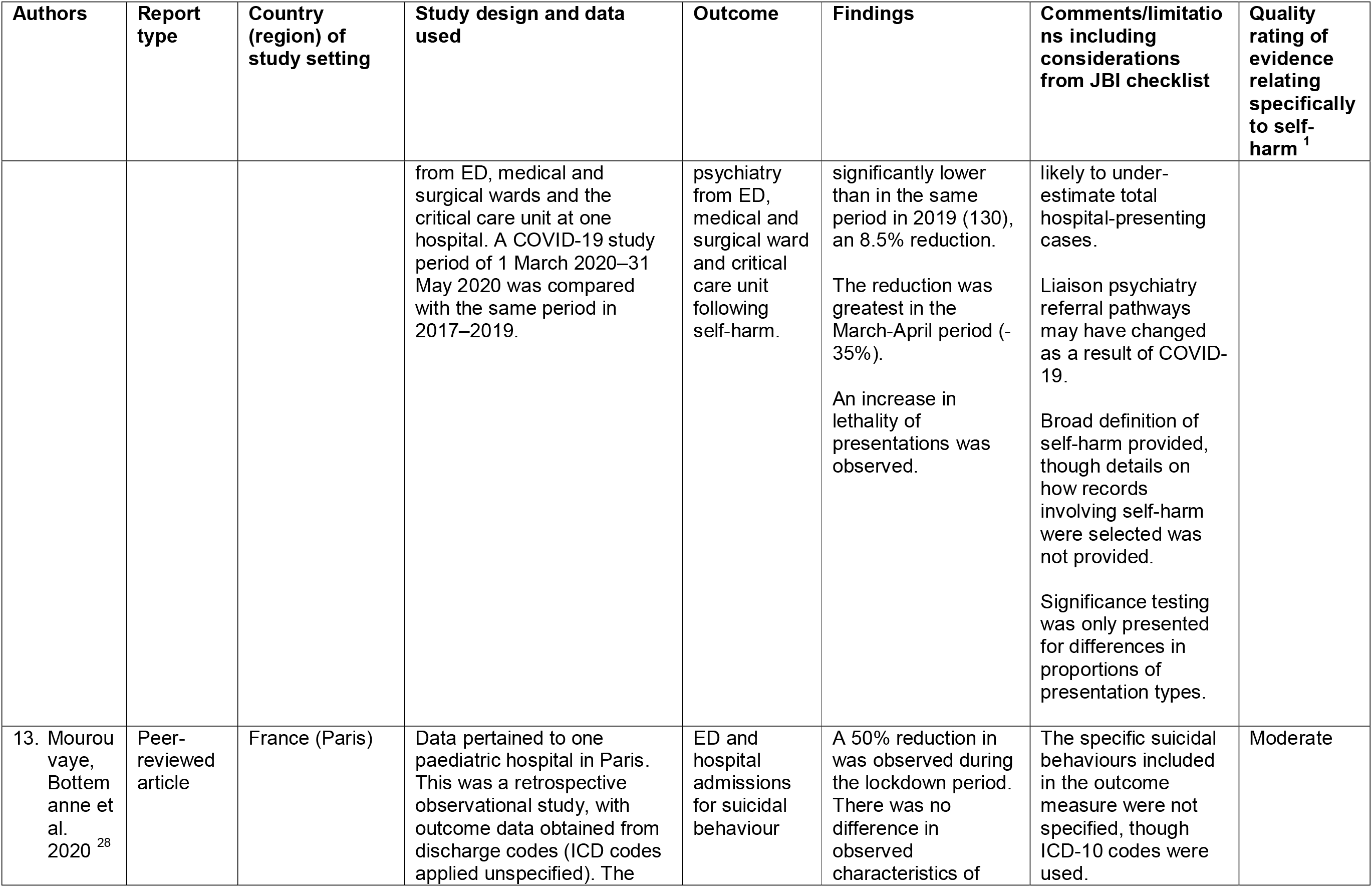
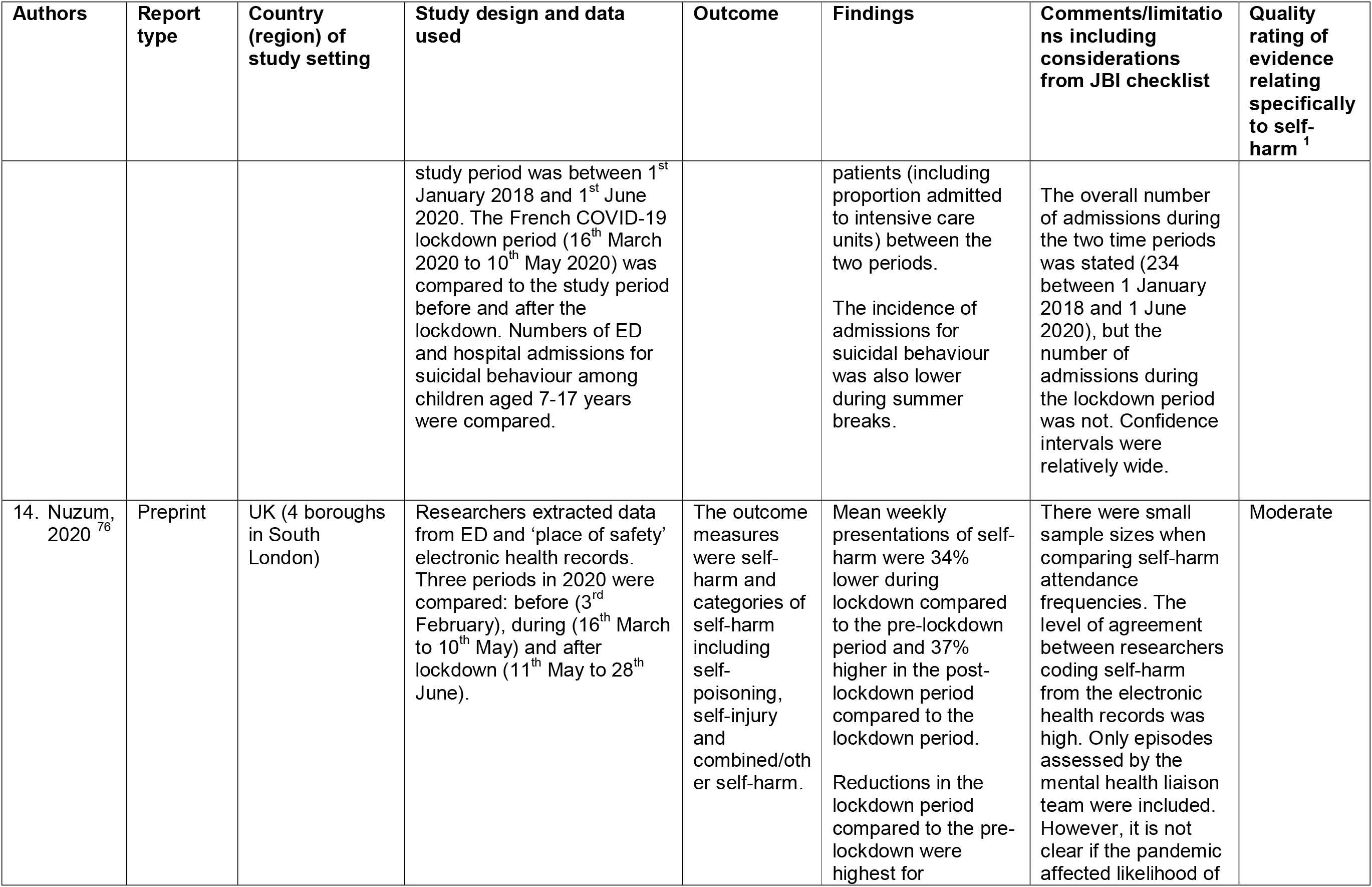
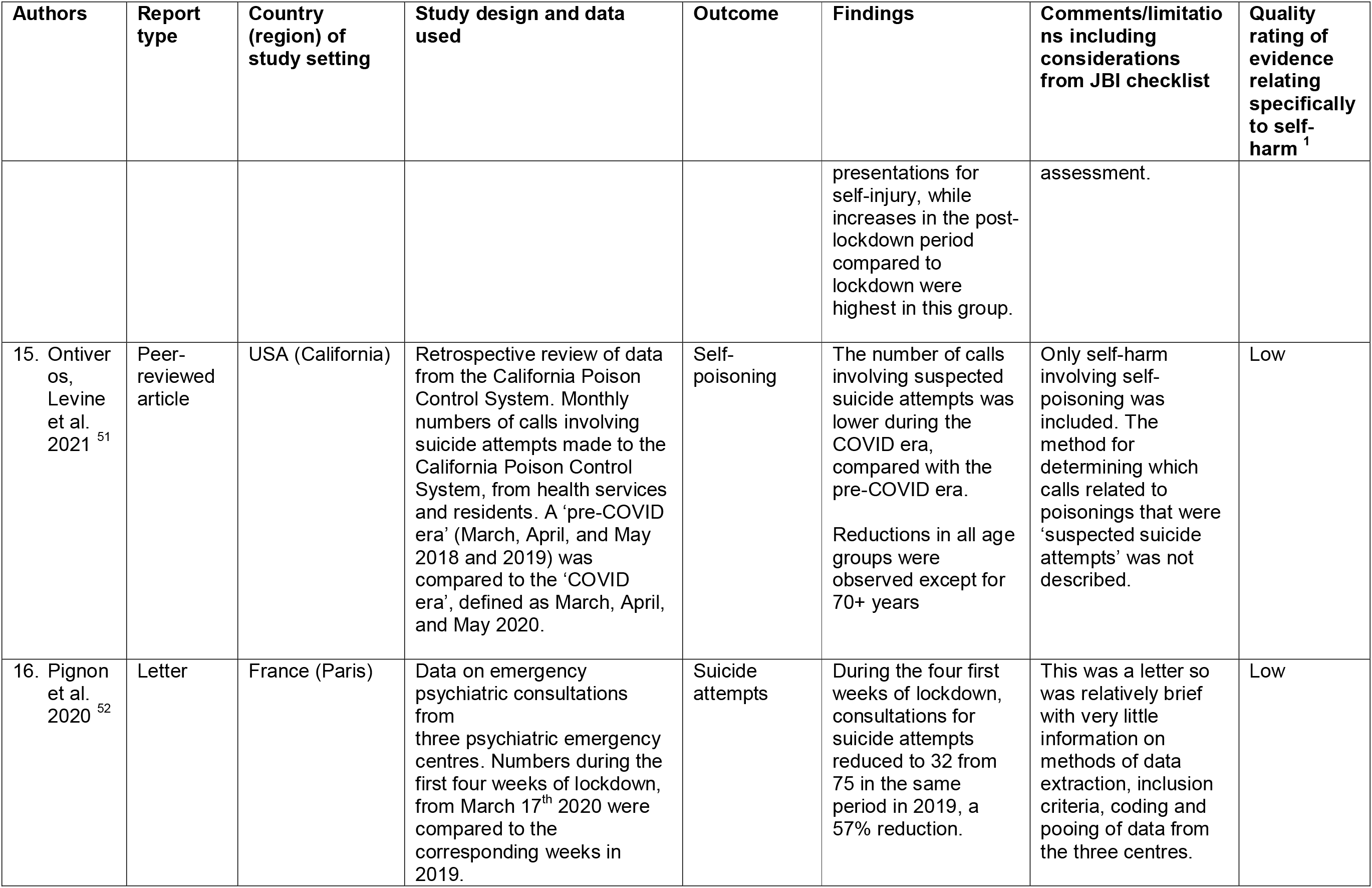
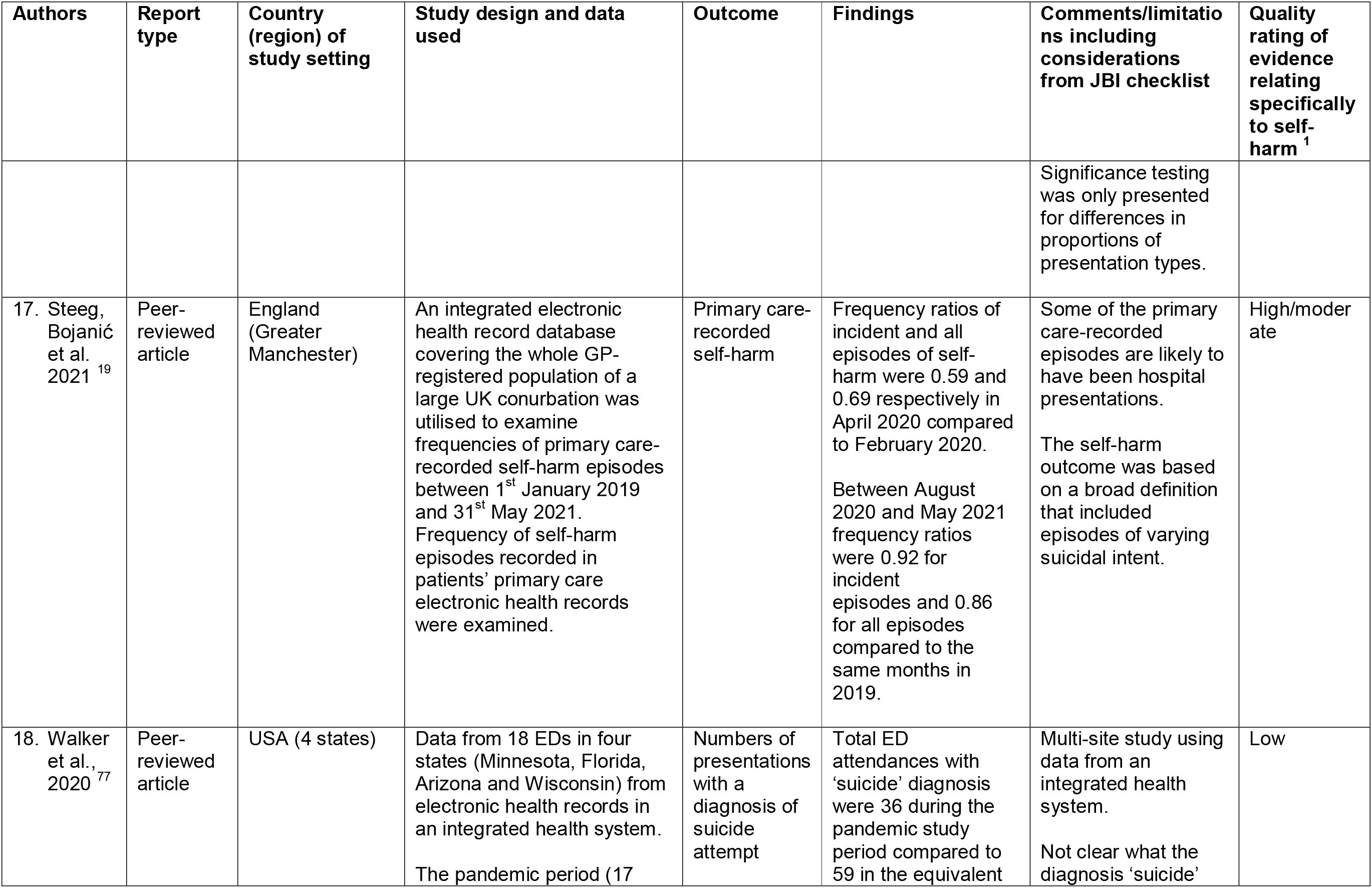
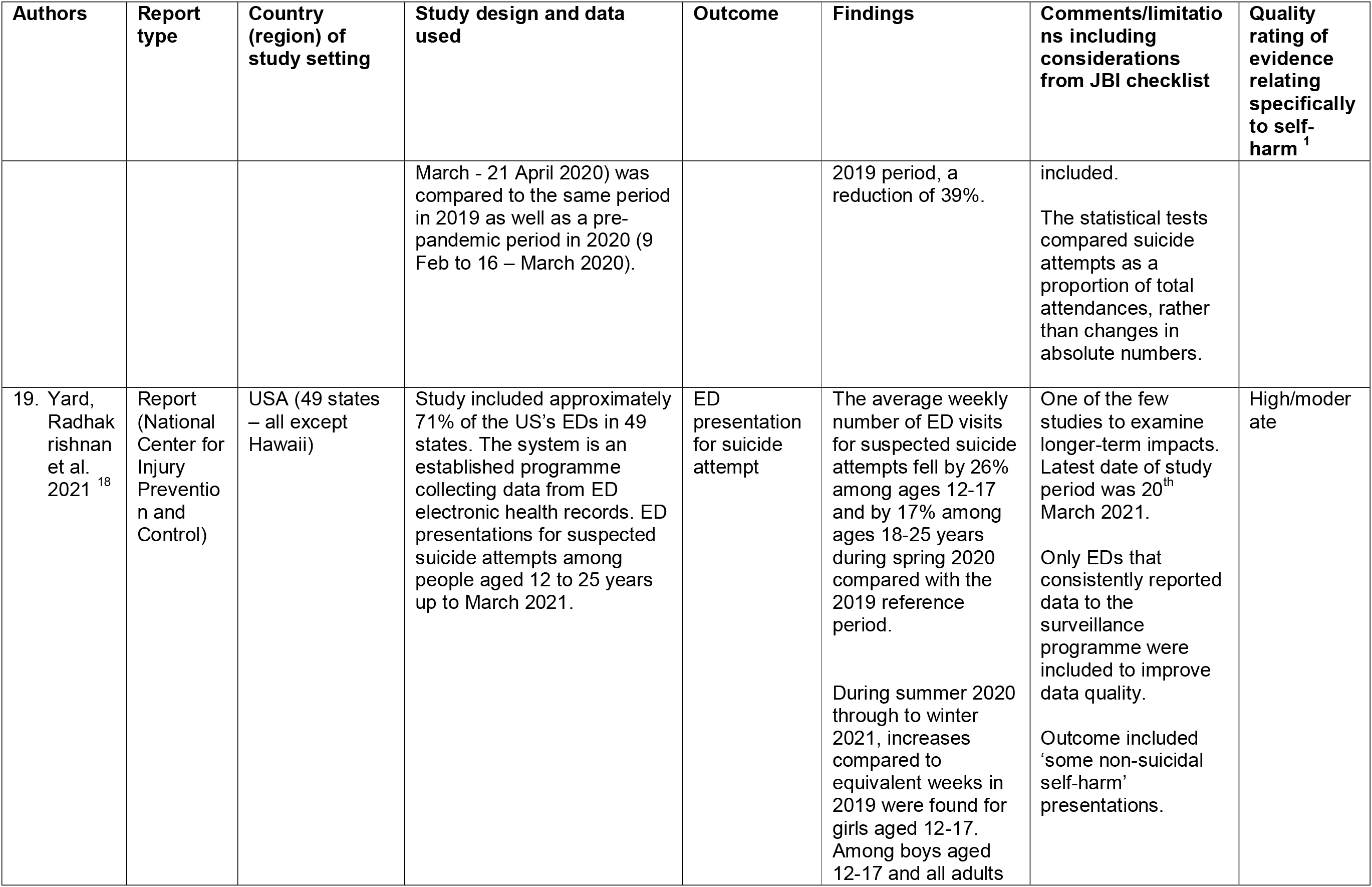
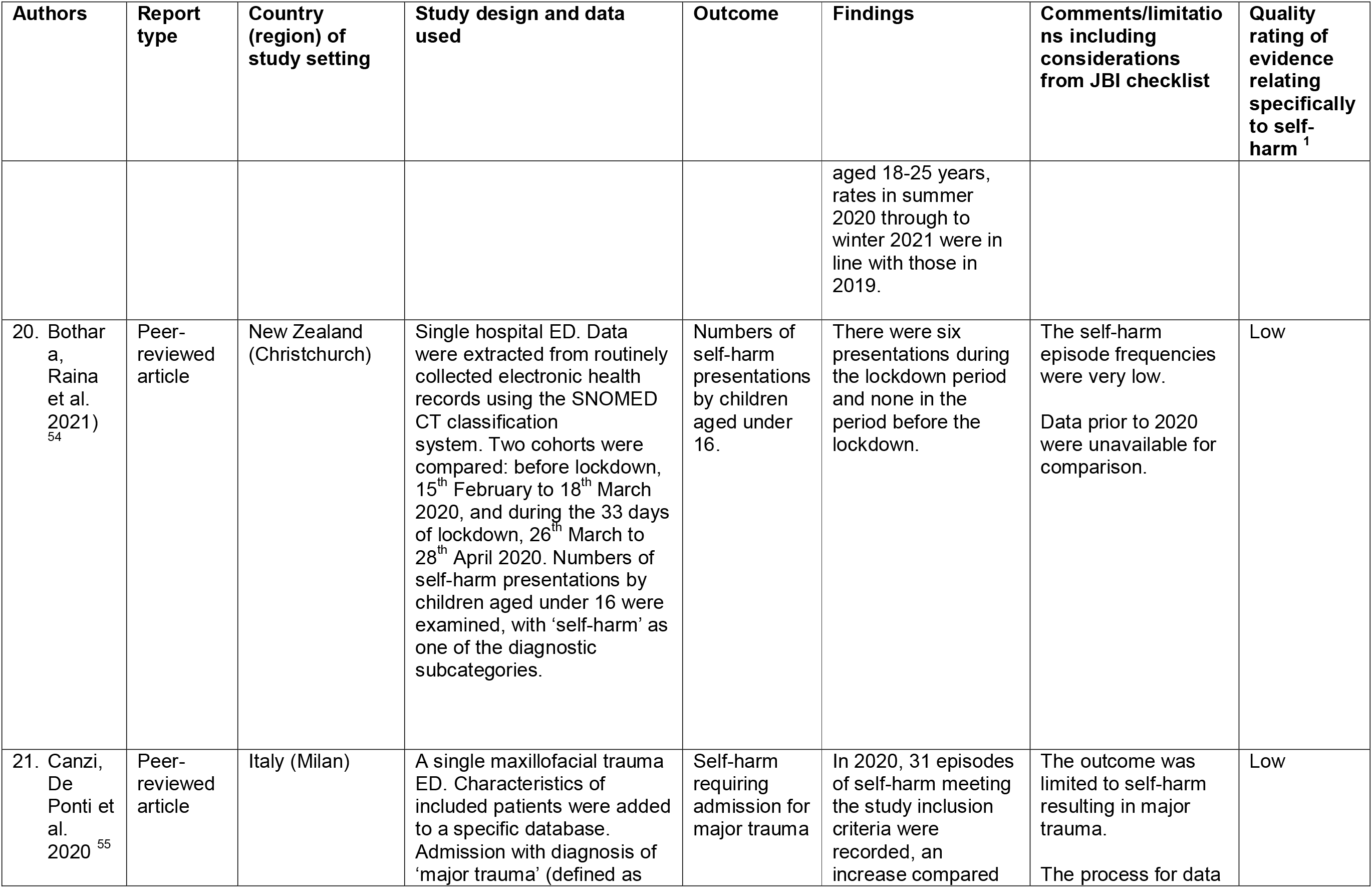
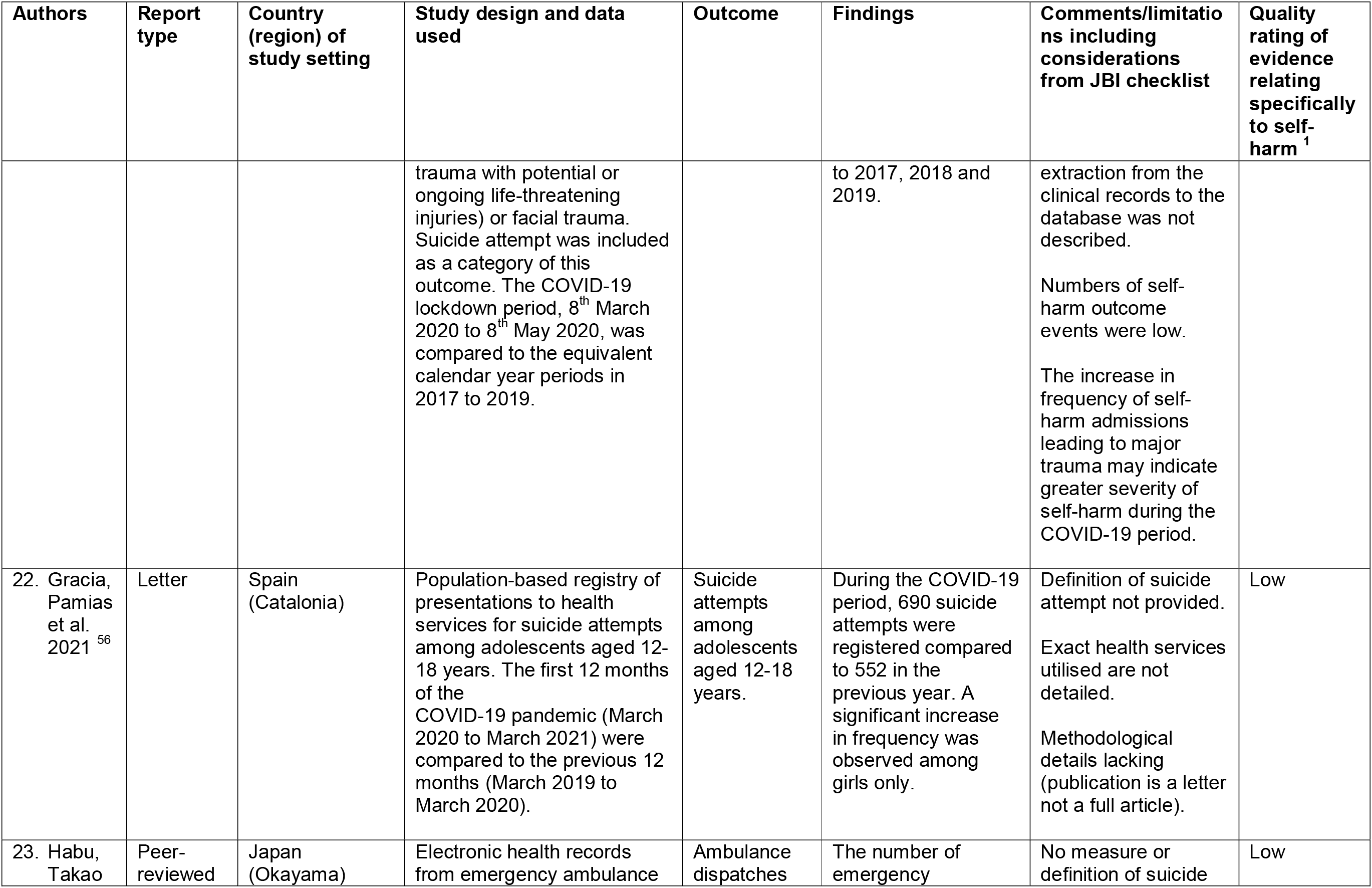
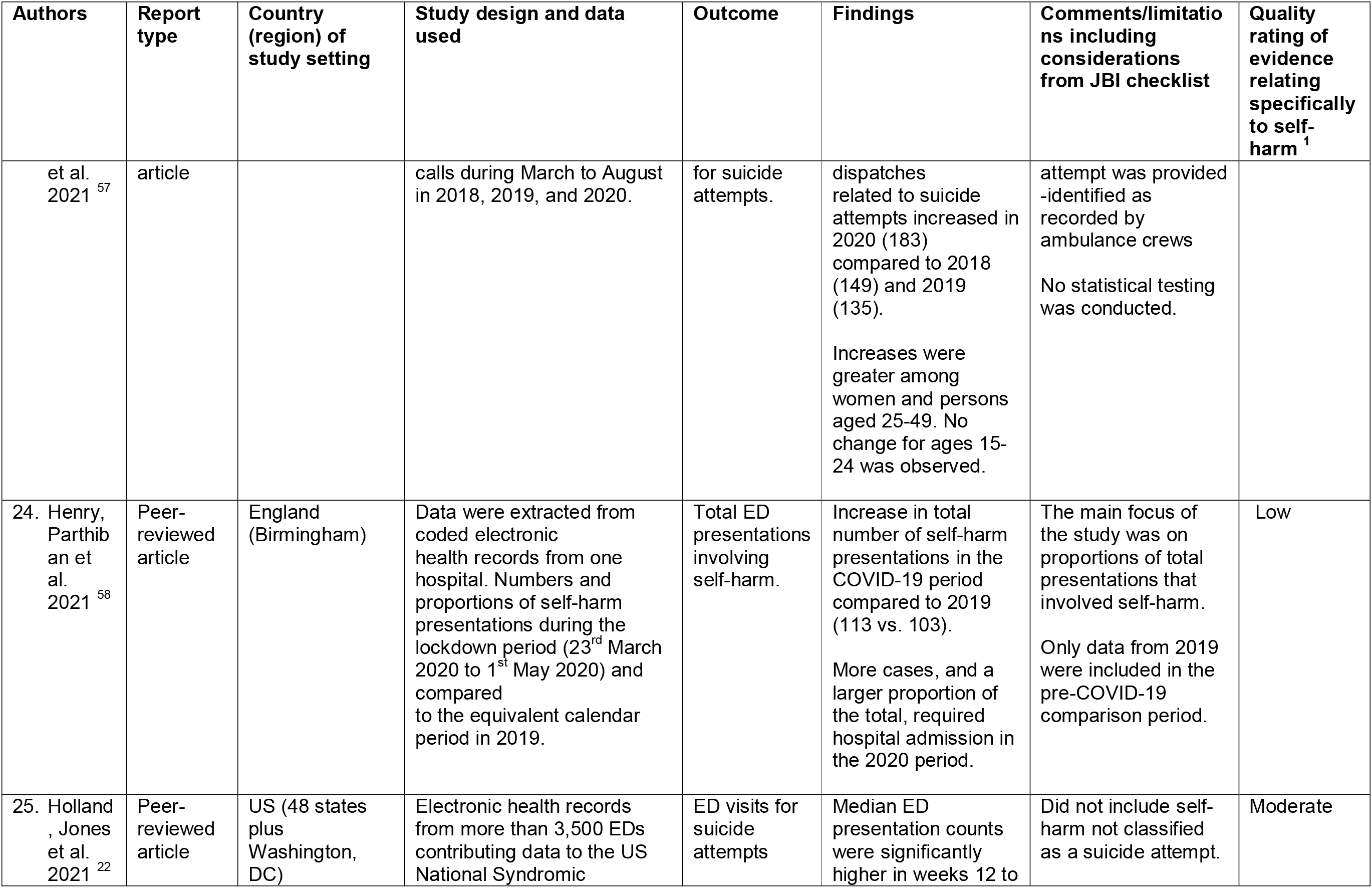
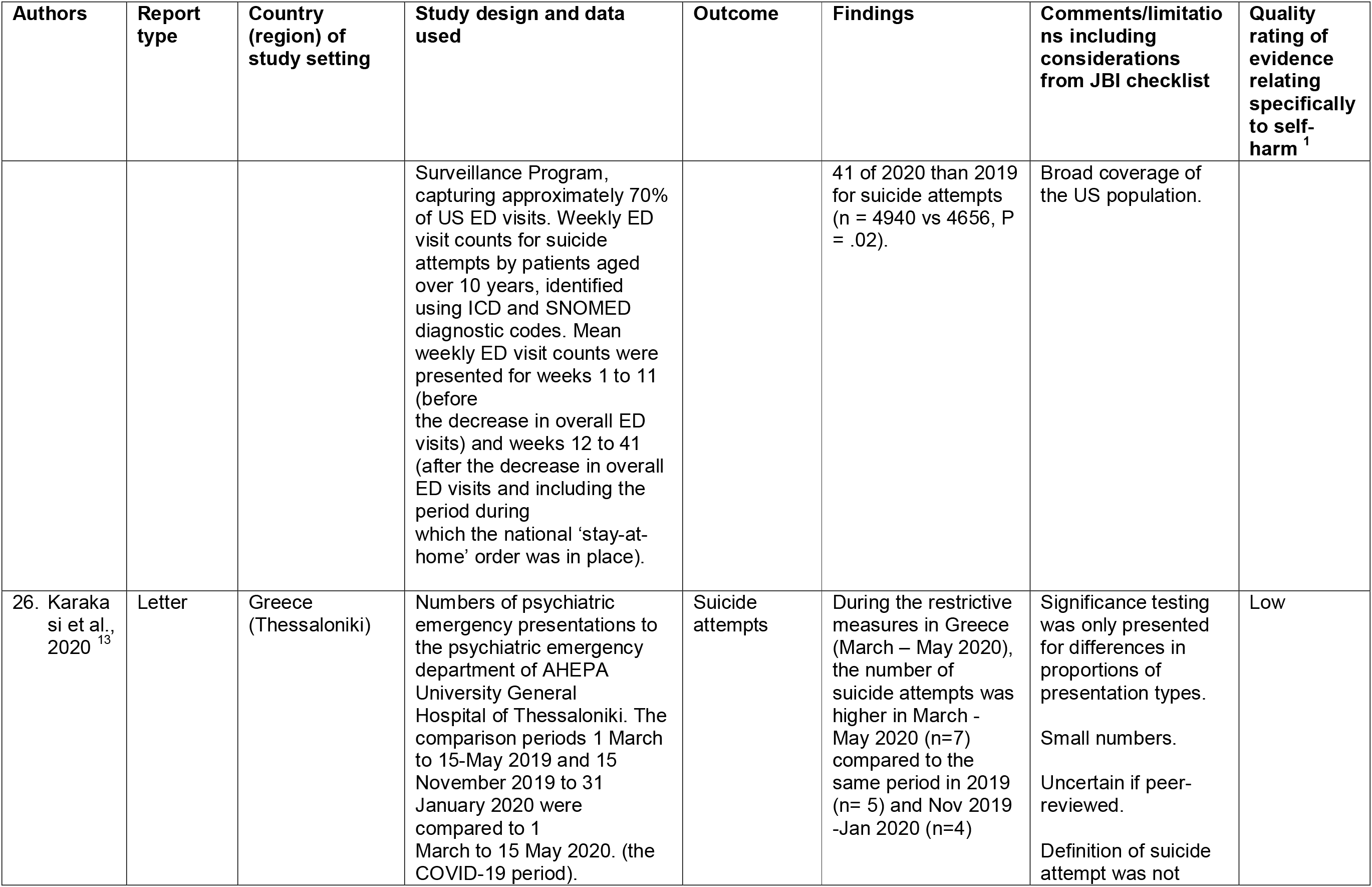
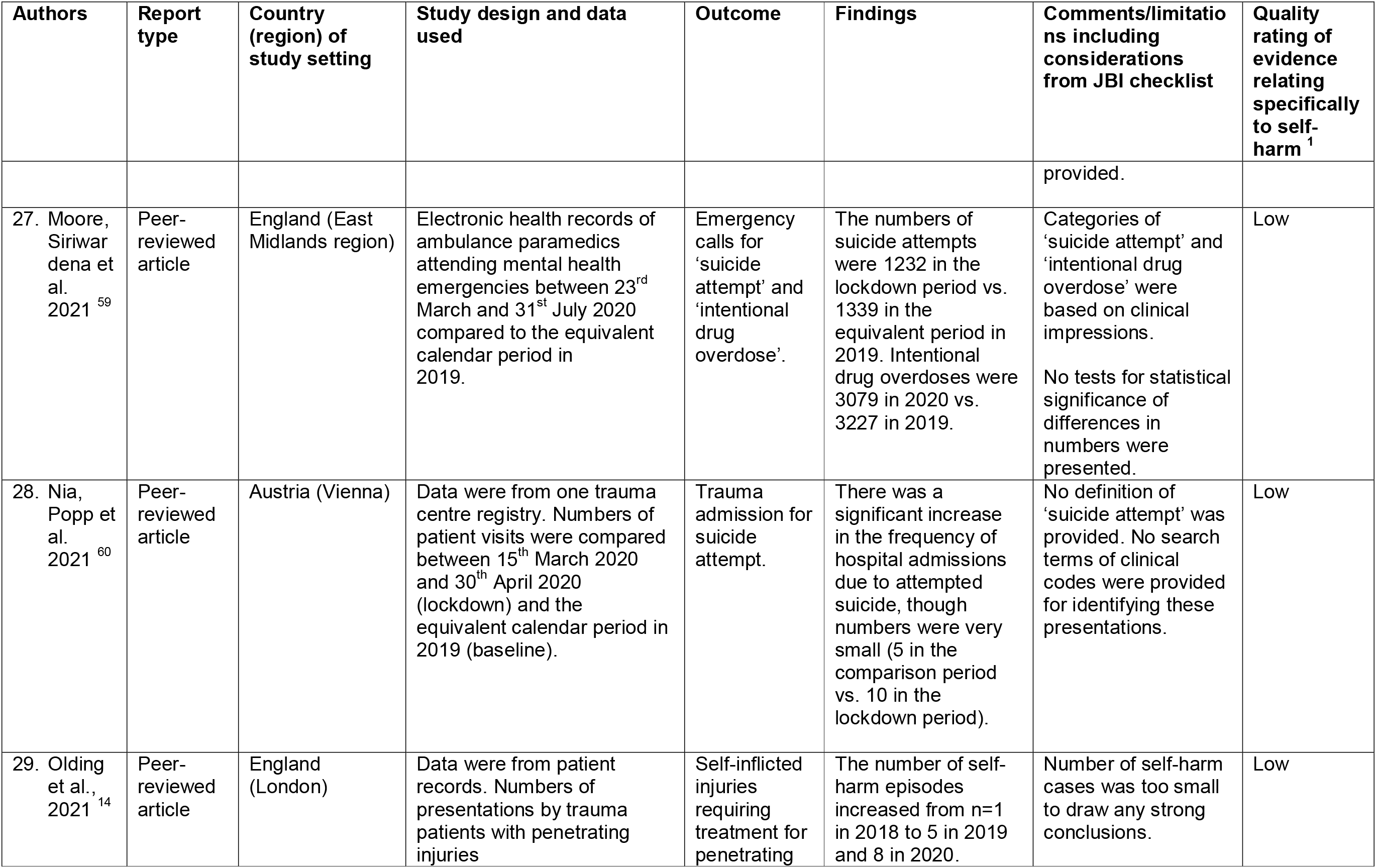
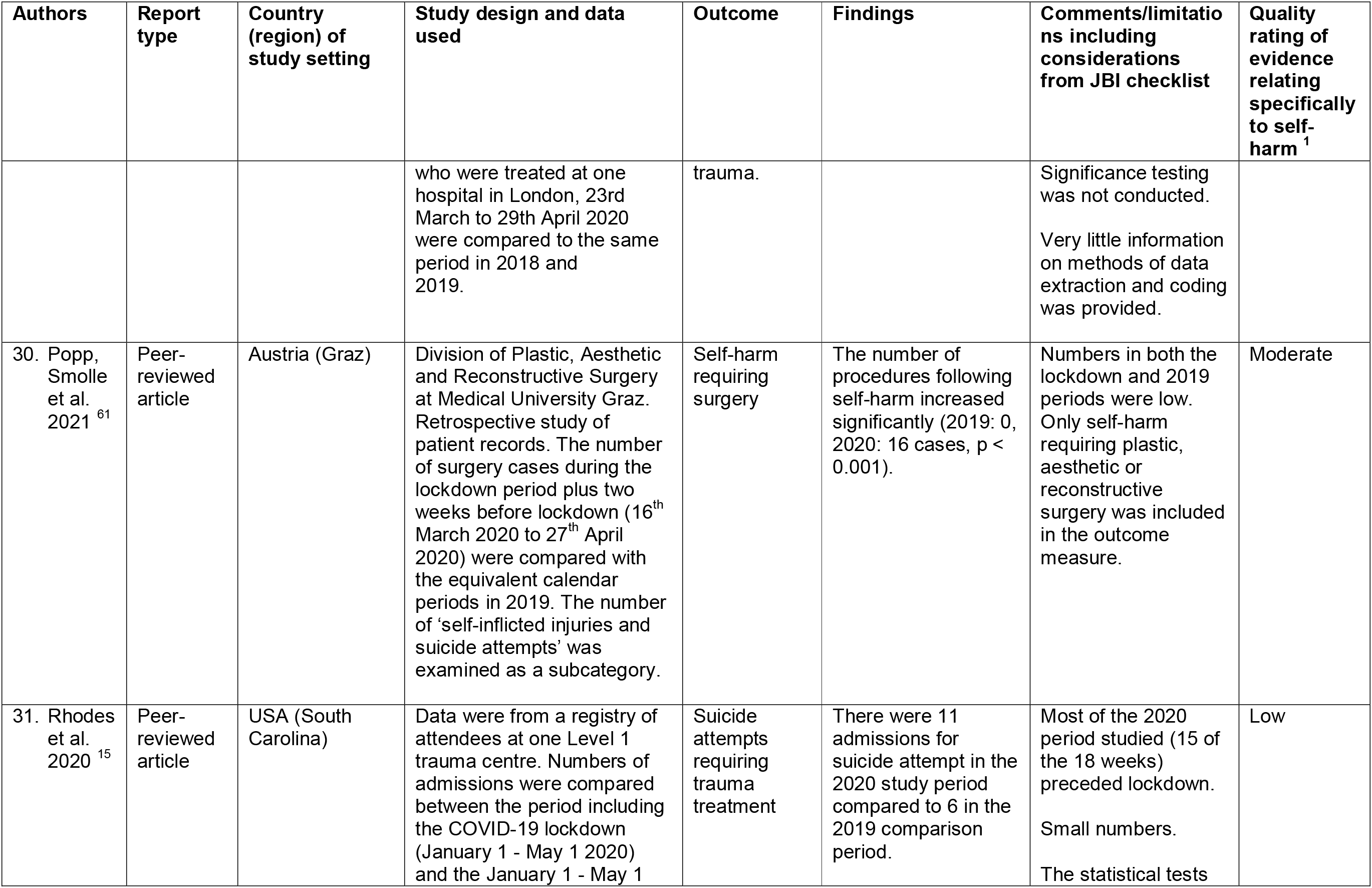
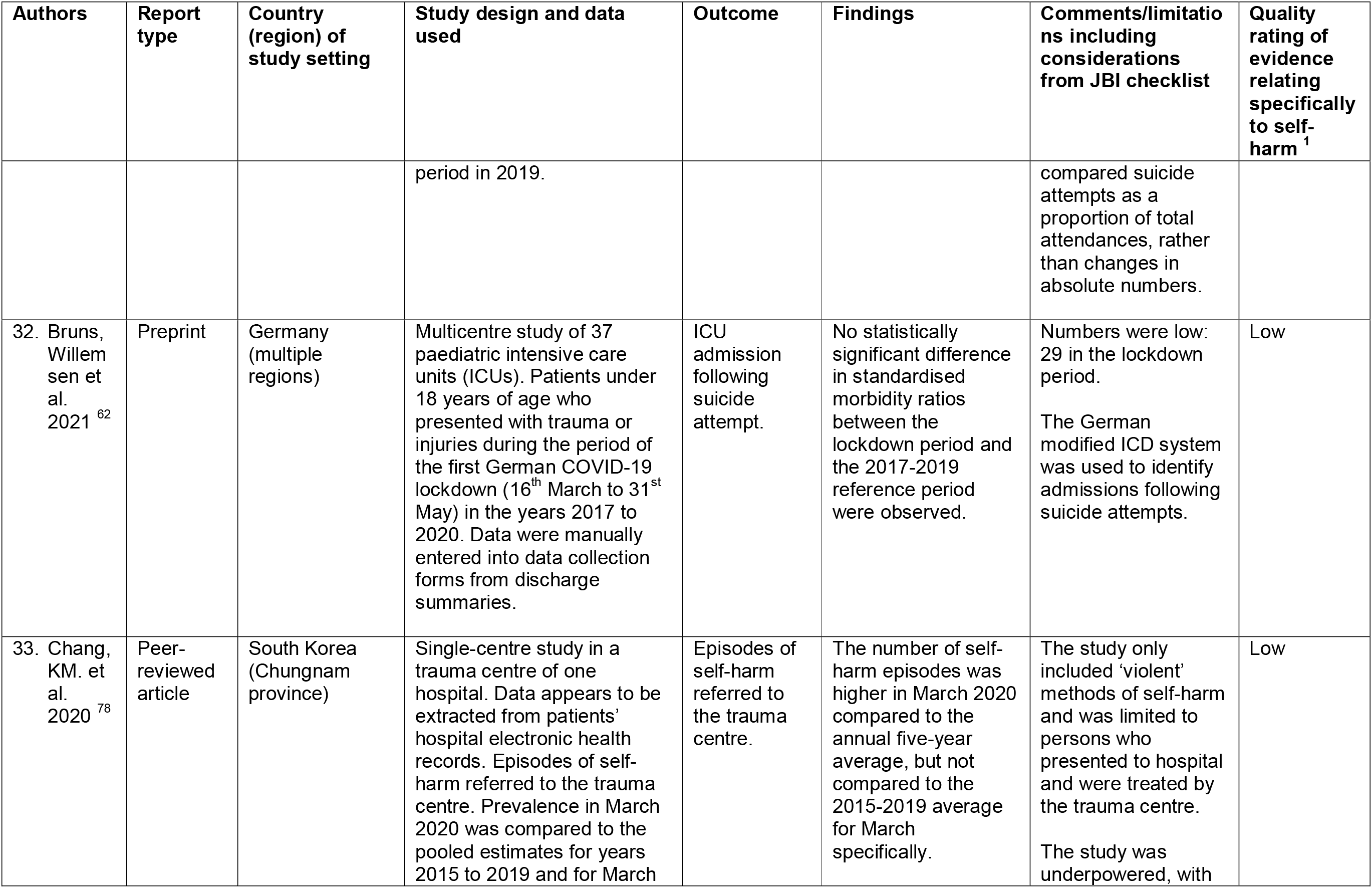
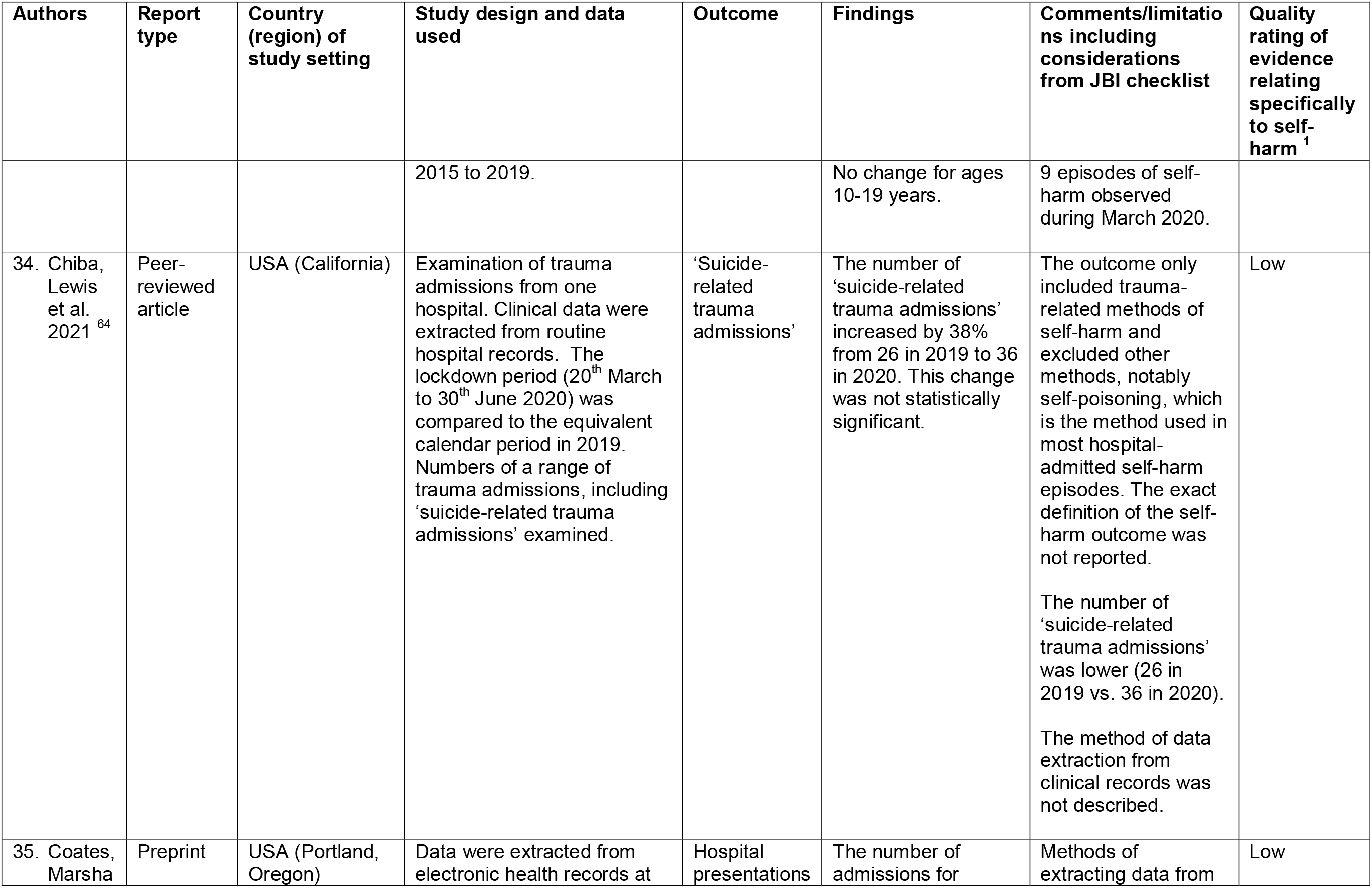
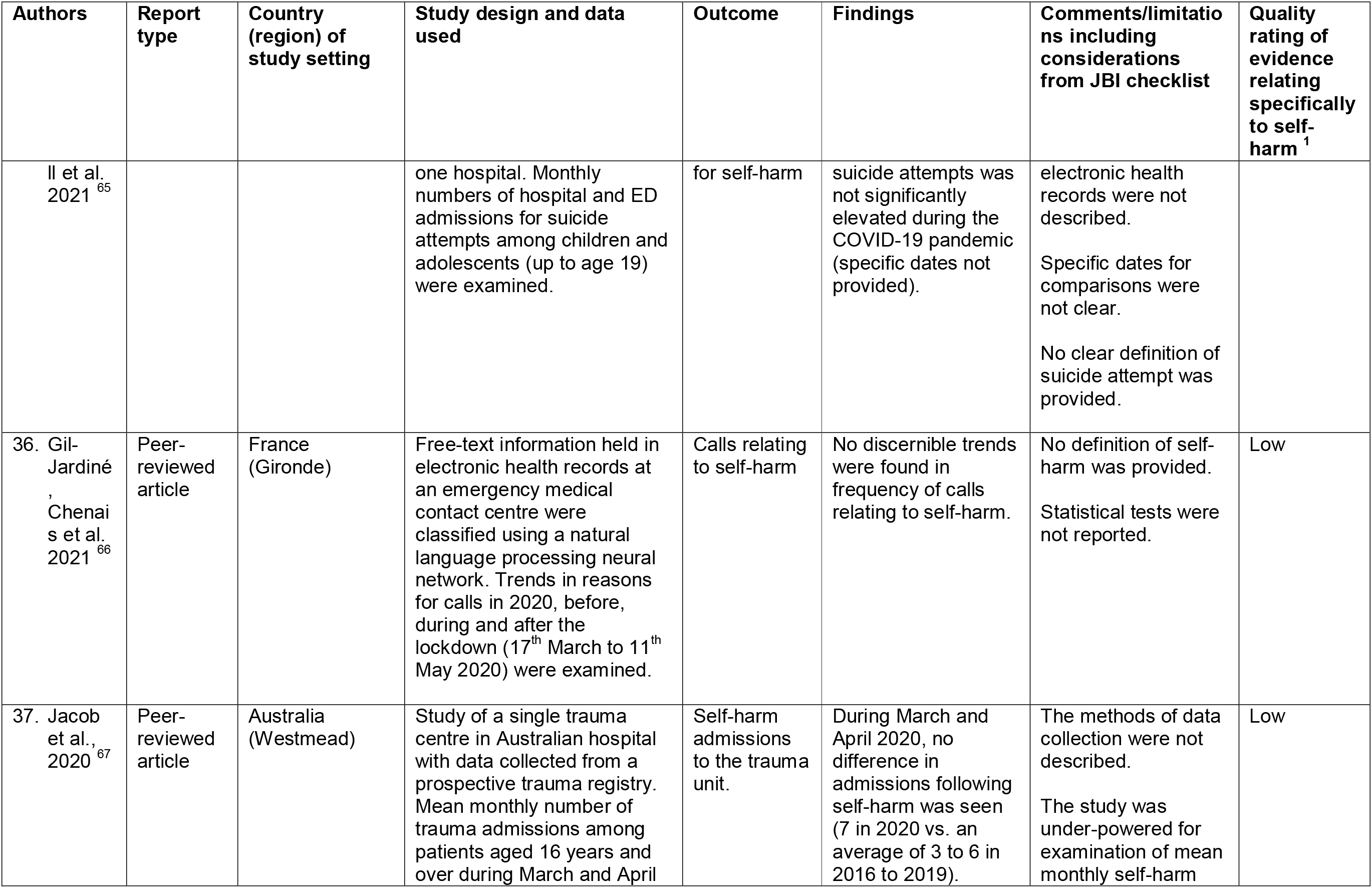
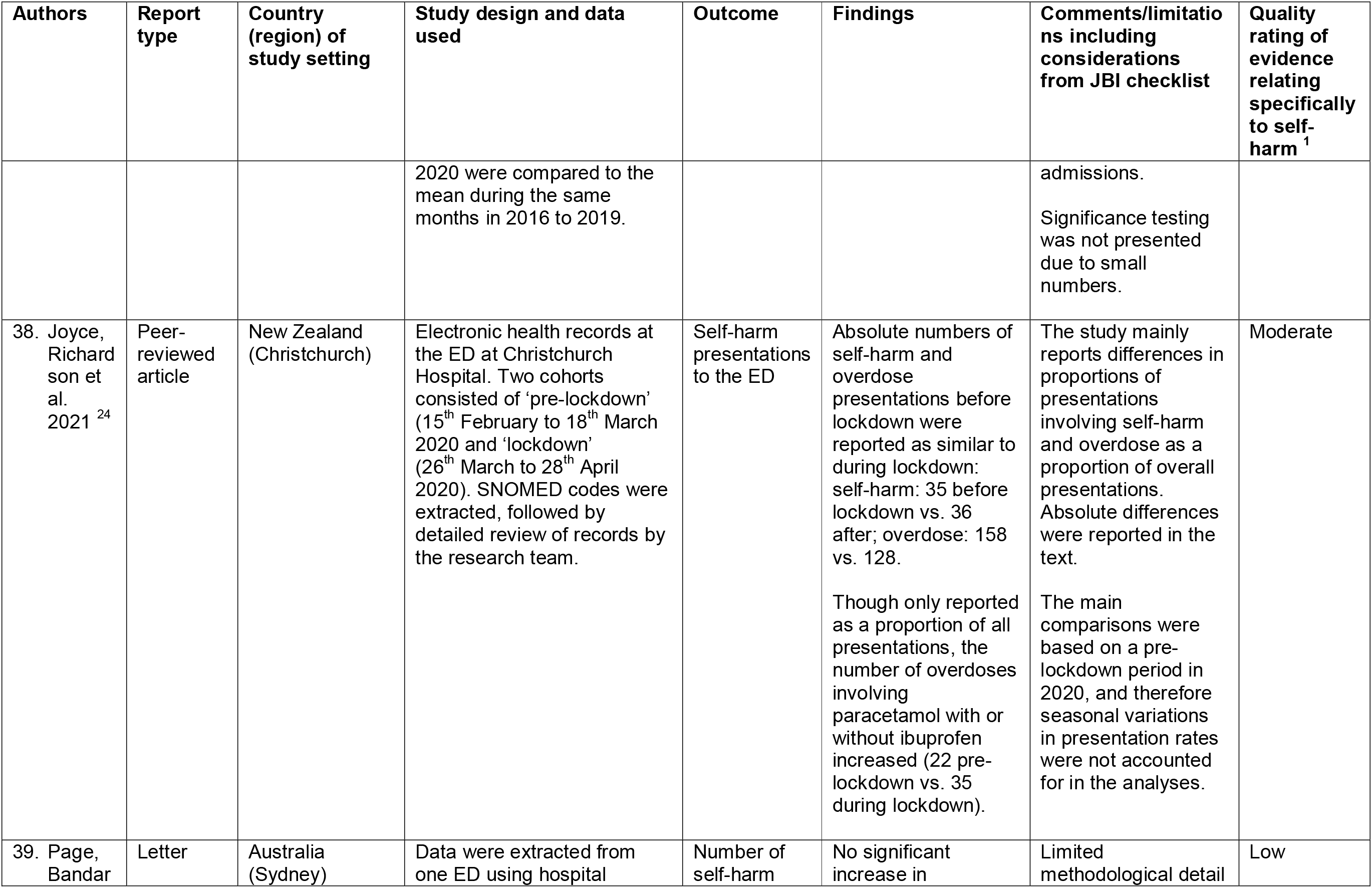
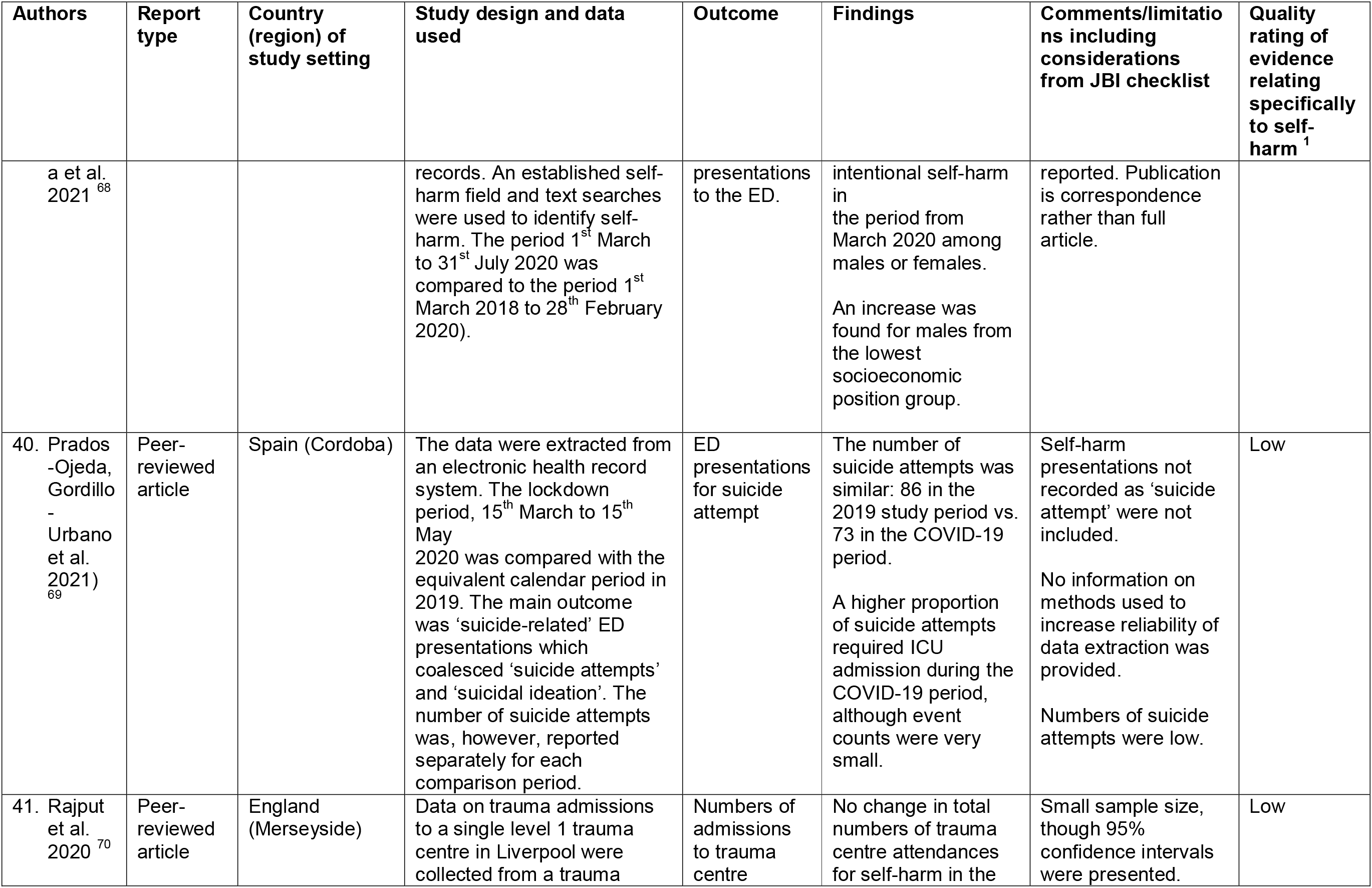
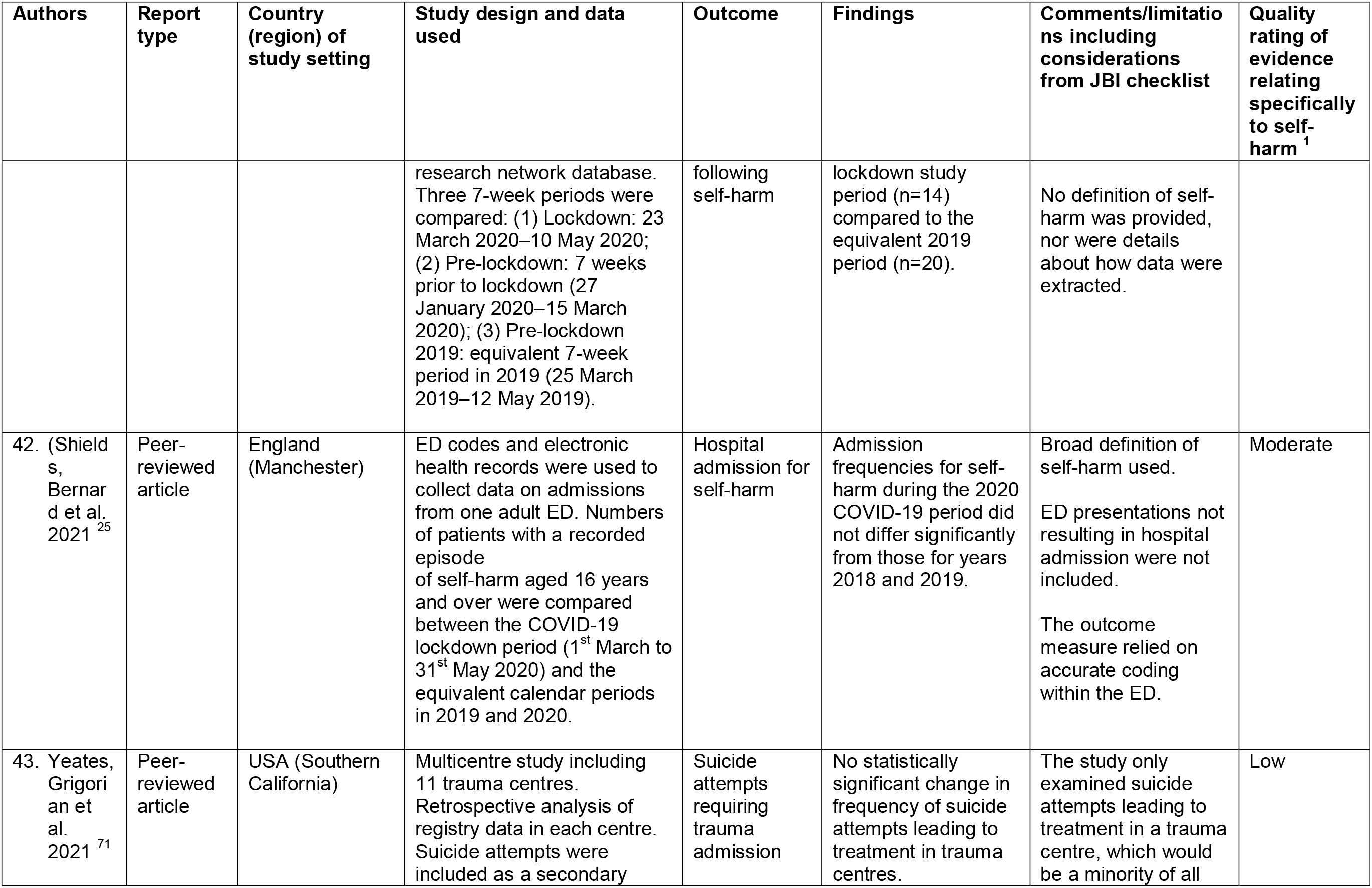
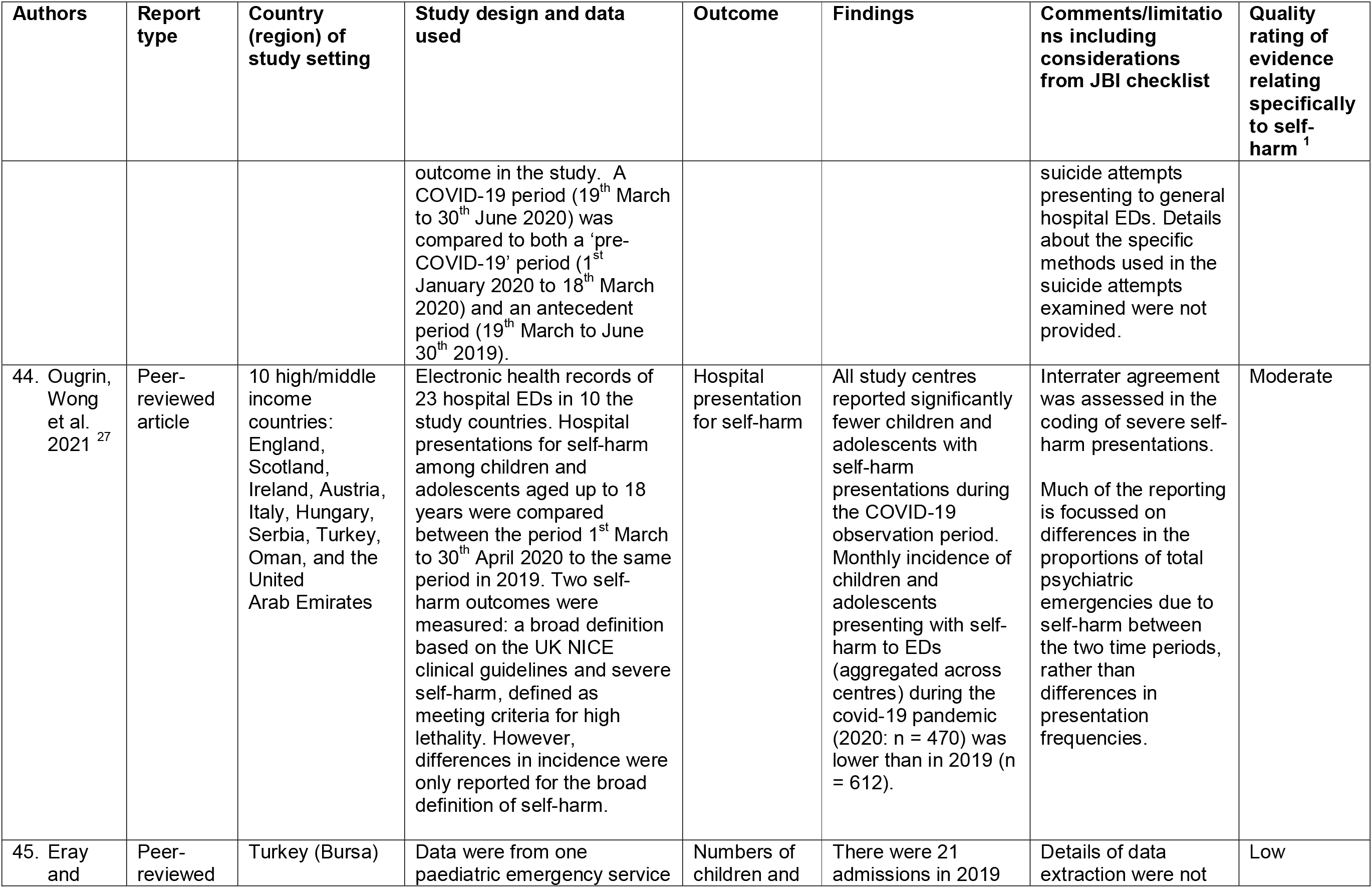
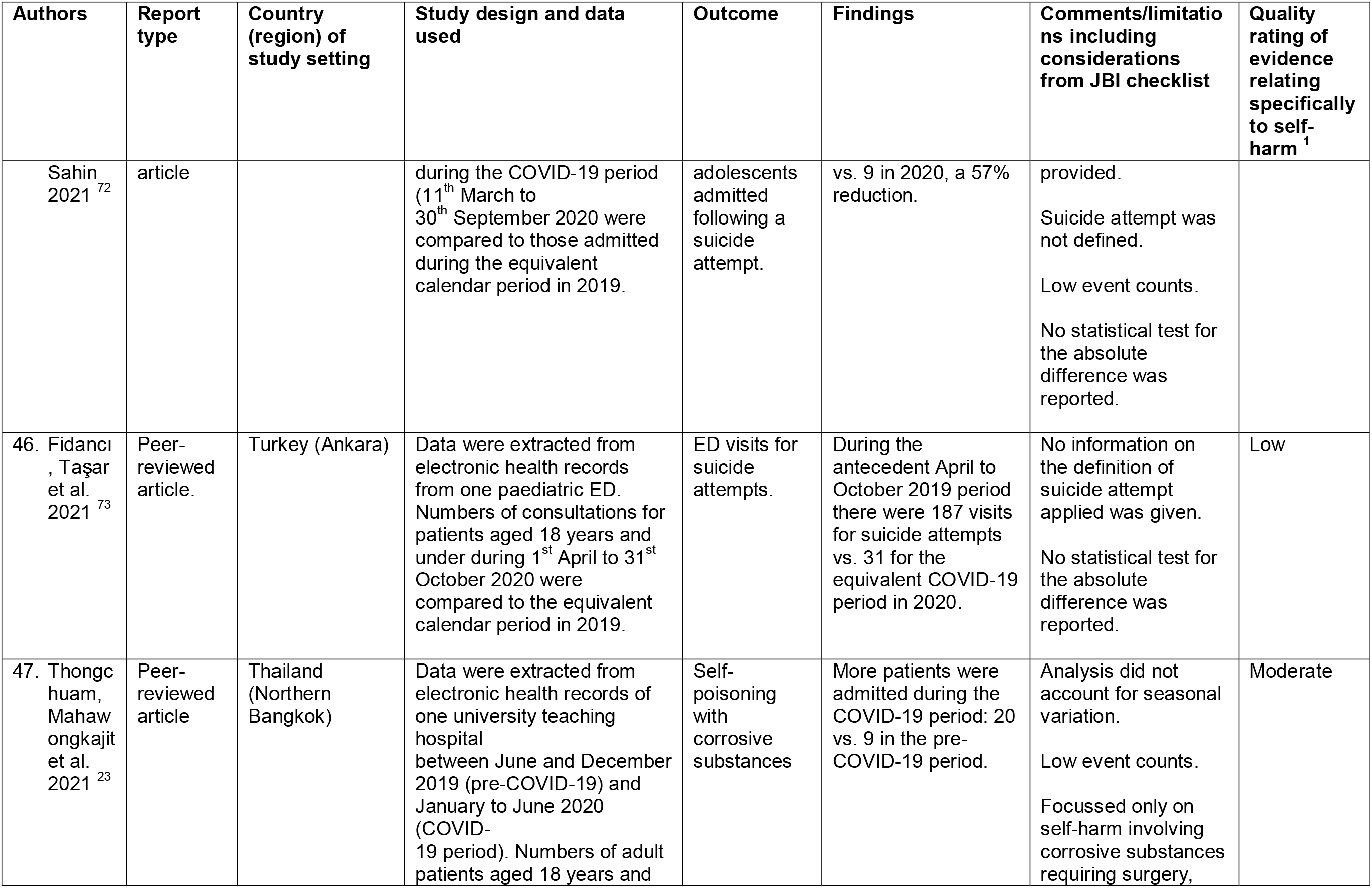
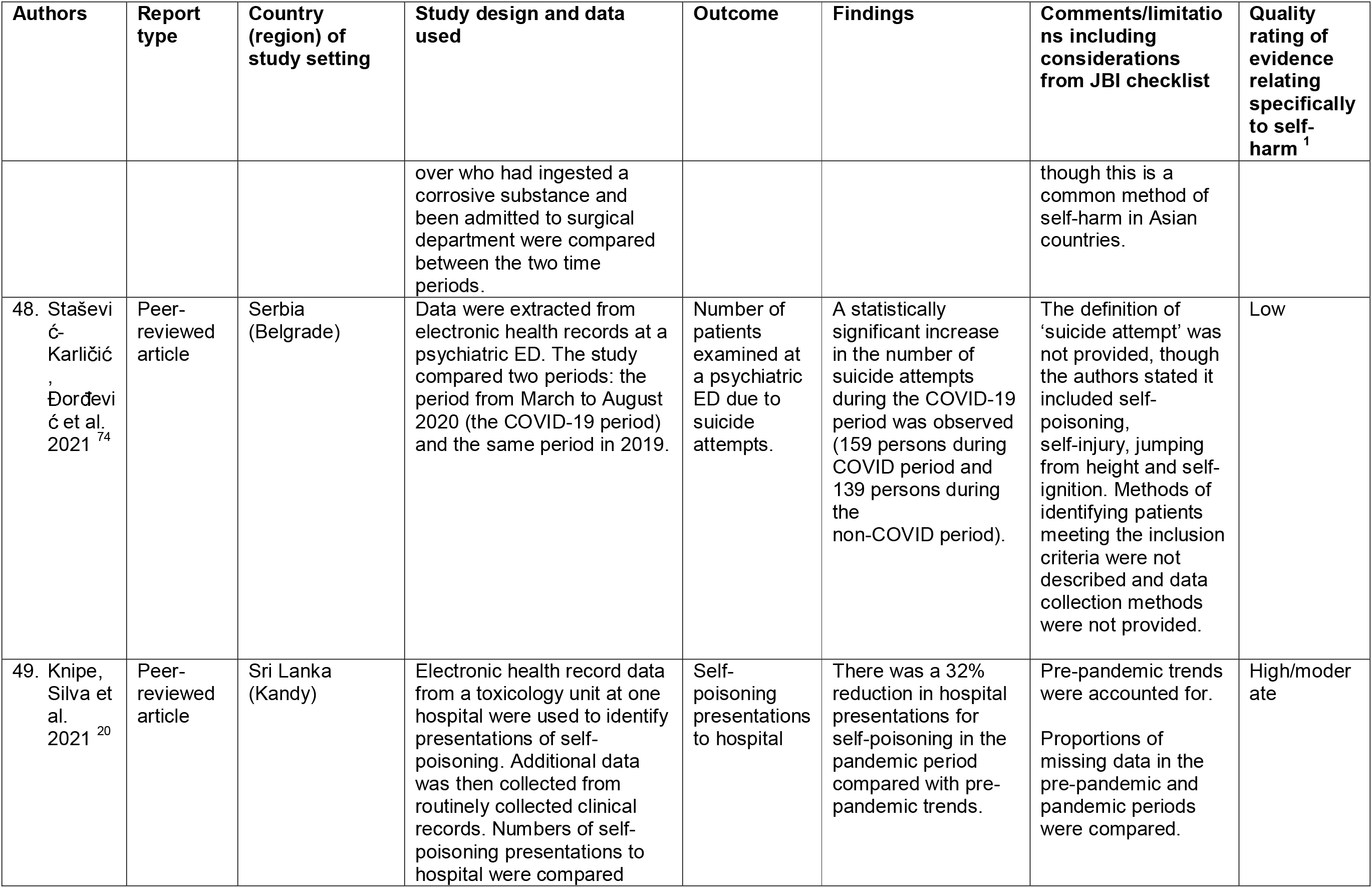
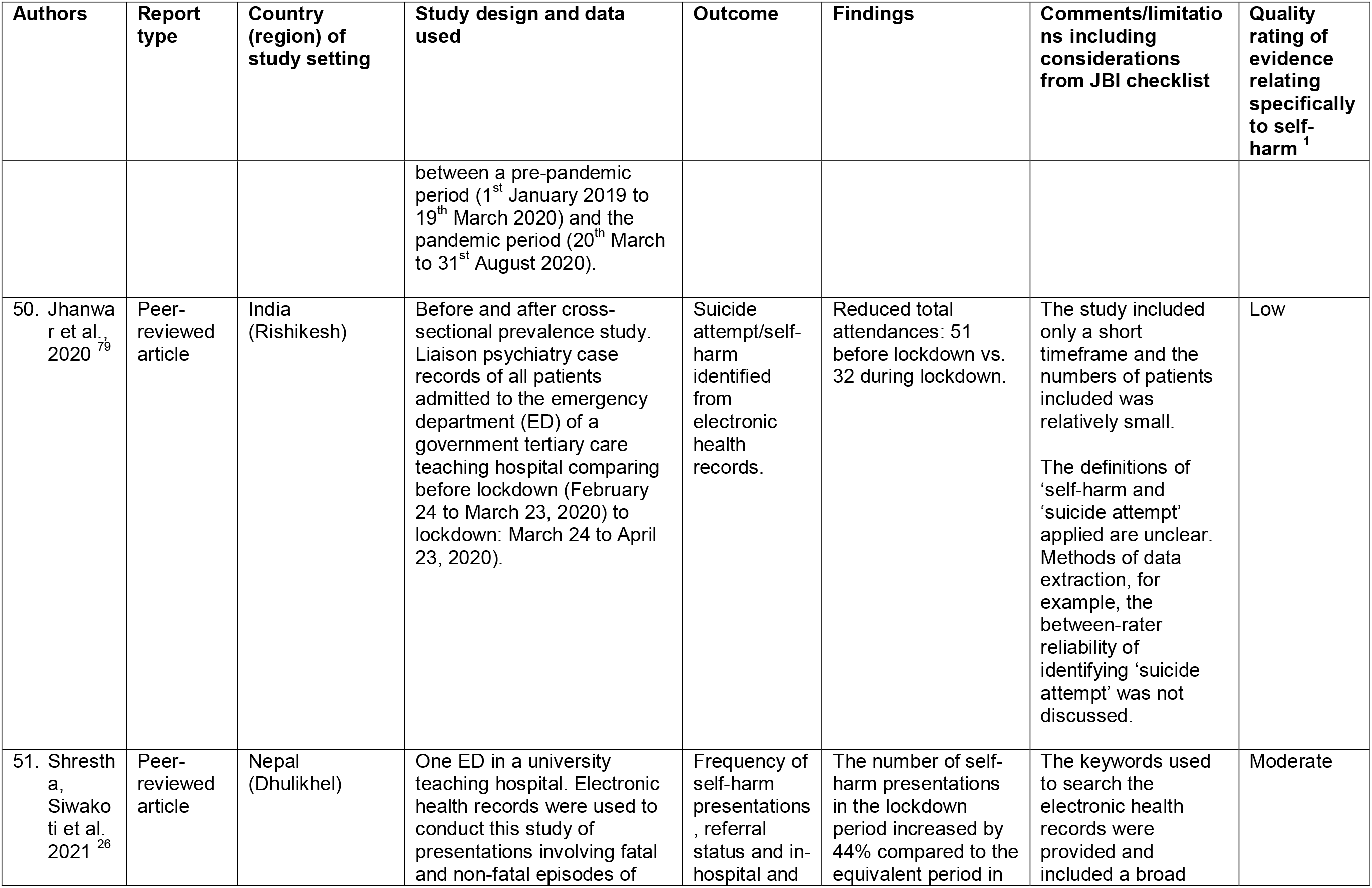
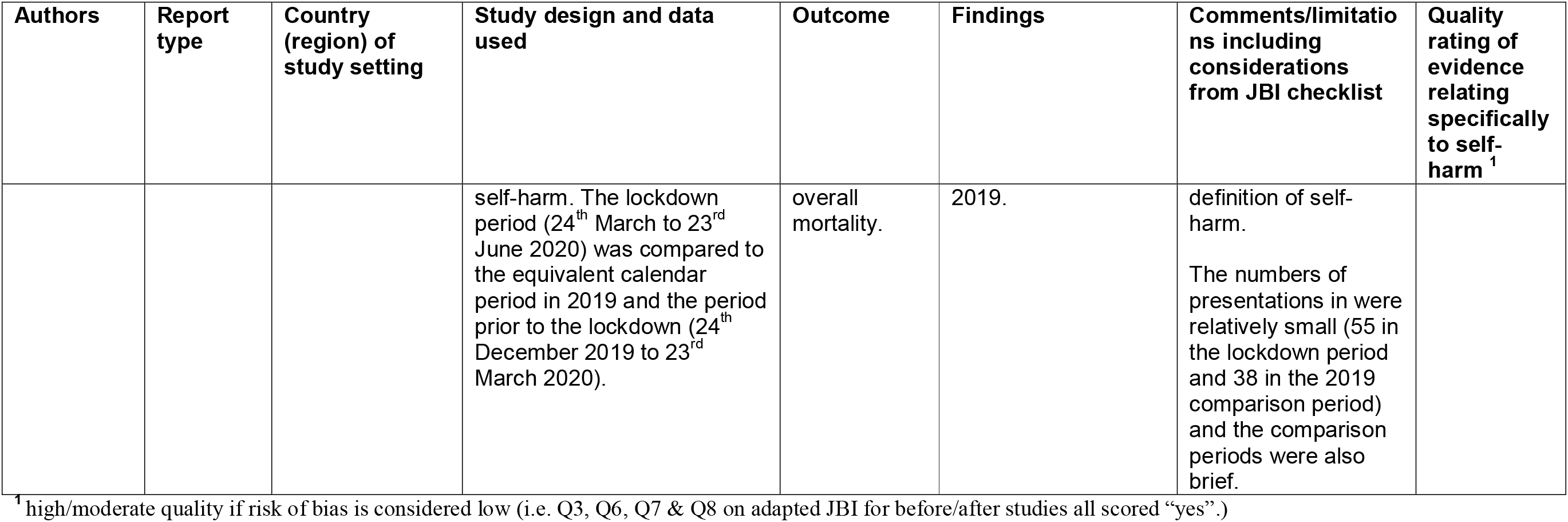
Details of included studies: 1^st^ Jan. 2020 to 7^th^ Sep. 2021

## Supplement 2: Search strategies for “The impact of the COVID-19 pandemic on self-harm and suicidal behaviour: update of living systematic review” and associated publications

### Scopus

TITLE-ABS-KEY(“selfharm*” OR “self harm*” OR “self-harm*” OR “self injur*” OR “selfinjur*” OR “self-injur*” OR “selfmutilat*” OR “self mutilat*” OR “self-mutilat*” OR “suicid*” OR “parasuicid*” OR “suicide” OR “suicidal ideation” OR “attempt* suicide” OR “suicide attempt*” OR “drug overdose” OR “selfpoisoning” OR “self poisoning” OR “self-poisoning” OR “self- injurious behavi*” OR “selfmutilation” OR “self mutilation” OR “self-mutilation” OR “automutilation” OR “suicidal behavi*” OR “selfdestructive behavi*” OR “self destructive behavi*” OR “self-destructive behavi*” OR “selfimmolat*” OR “self-immolat*” OR “self immolat*” OR “cutt*” OR “headbang” OR “head-bang” OR “head bang” OR “overdose” OR “selfinflict*” OR “self-inflict*” OR “self inflict*” OR “hopelessness” OR “powerlessness” OR “helplessness” OR “negative attitude*” OR “emotional negativism” OR “pessimism” OR “depress*” OR “hopelessness depression” OR “passivity” OR “sad-affect” OR “sadness” OR “decreased affect” OR “cognitive rigidity” OR “suicidality” OR “suicide ideation”) AND TITLE-ABS-KEY(“nCoV” OR “HCoV” OR “covid 19” OR “covid-19” OR “covid19” OR “coronavirus” OR “19 ncov” OR “19-ncov” OR “2019 ncov” OR “2019-ncov” OR “2019ncov” OR “n-cov” OR “ncov” OR “coronavirus disease*” OR “sars-cov-2” OR “sars cov 2” OR “sars-cov 2” OR “mers-cov” OR “mers cov”) AND PUBYEAR > 2020

*The filter ‘PUBYEAR > 2020’ corresponds to the 2021 version of this search, in previous years we used ‘PUBYEAR > 2018’ and ‘PUBYEAR > 2019’ respectively*

### Medline via PubMed

((mental health[TIAB] OR selfharm*[TIAB] OR self-harm*[TIAB] OR selfinjur*[TIAB] OR self- injur*[TIAB] OR selfmutilat*[TIAB] OR self-mutilat*[TIAB] OR suicid*[TIAB] OR parasuicid*[TIAB) OR (suicide[TIAB] OR suicidal ideation[TIAB] OR attempted suicide[TIAB]) OR (drug overdose[TIAB] OR self?poisoning[TIAB]) OR (self-injurious behavio?r[TIAB] OR self?mutilation[TIAB] OR automutilation[TIAB] OR suicidal behavio?r[TIAB] OR self?destructive behavio?r[TIAB] OR self?immolation[TIAB])) OR (cutt*[TIAB] OR head?bang[TIAB] OR overdose[TIAB] OR self?immolat*[TIAB] OR self?inflict*[TIAB]) OR (hopelessness[TIAB] OR powerlessness[TIAB] OR helplessness[TIAB] OR negative attitude$[TIAB] OR emotional negativism[TIAB] OR pessimism[TIAB] OR depress*[TIAB] OR hopelessness depression[TIAB] OR passivity[TIAB] OR sad-affect[TIAB] OR sadness[TIAB] OR decreased affect[TIAB] OR cognitive rigidity[TIAB] OR suicidality[TIAB] OR suicide ideation[TIAB]))) AND ((coronavirus disease?19[TIAB] OR sars?cov?2[TIAB] OR mers?cov[TIAB]) OR (19?ncov[TIAB] OR 2019?ncov[TIAB] OR n?cov[TIAB]) OR (\“severe acute respiratory syndrome coronavirus 2\” [Supplementary Concept] OR \“COVID-19\” [Supplementary Concept] OR COVID-19 [tw] OR COVID 2019 [tw] OR coronavirus [tw] OR nCoV[TIAB] OR HCoV))

### Psy- and SocArXiv (both same query)

“(mental health OR selfharm* OR self-harm* OR selfinjur* OR self-injur* OR selfmutilat* OR self-mutilat* OR suicid* OR parasuicid* OR suicide OR suicidal ideation OR attempted suicide OR drug overdose OR self?poisoning OR self-injurious behavio?r OR self?mutilation OR automutilation OR suicidal behavio?r OR self?destructive behavio?r OR self?immolation OR cutt* OR head?bang OR overdose OR self?immolat* OR self?inflict* OR hopelessness OR powerlessness OR helplessness OR negative attitude OR emotional negativism OR pessimism OR depress* OR hopelessness depression OR passivity OR sad-affect OR sadness OR decreased affect OR cognitive rigidity OR suicidality OR suicide ideation) AND (coronavirus disease?19 OR sars?cov?2 OR mers?cov OR 19?ncov OR 2019?ncov OR n?cov OR COVID-19 OR COVID 2019 OR coronavirus OR nCoV OR HCoV)”

### No date/content filters applied

Med and BioRxiv, WHO Covid-19 database

We directly retrieve ALL new publications related to Covid-19 from these sources, see http://connect.biorxiv.org/relate/content/181 for Bio- MedRxiv Covid feed and https://search.bvsalud.org/global-literature-on-novel-coronavirus-2019-ncov/ for WHO on a daily basis. There is no date filter, we retrieve each new report as it becomes available. We then apply our systematic search to those results, as described here: McGuinness et al., (2020). medrxivr: Accessing and searching medRxiv and bioRxiv preprint data in R. Journal of Open Source Software, 5(54), 2651. https://doi.org/10.21105/joss.02651 The search strategy is below, please note that the syntax is RegularExpression, but it was designed to correspond to the PubMed query you see above, with new lines being joined with ‘OR’ statements. The only exception is the removal of the AND statement relating to COVID-19 itself, because those 3 sources only include COVID-specific information.

[Ss]elf[- ]?[Ii]njur(y|ious)[- ]?[Bb]ehaviou?r

[Ss]elf[- ]?([Mm]utilat¬[Ii]mmolat)(ion|ed)

[Aa]uto[- ]?[Mm]utilat(ion|ed) [Ss]uicidal[- ]?[Bb]ehaviou?r

[Ss]elf[- ]?[Dd]estructive)[- ]?[Bb]ehaviou?r [Ss]uicide

[Aa]ttempted[- ]?[Ss]uicide [Ss]uicidal[- ]?[Ii]deation [Ss]elf[- ]?[Hh]arm

[Ss]elf[- ]?[Mm]utilat

[Ss]elf[- ]?[Ii]njur

[Pp]ara[ -]?[Ss]uicid

[Dd]rug[ -]?[Oo]verdose

[Ss]elf[- ]?[Pp]oison(ing|ed)

[Ss]elf[- ]?[iI]nflict

[Ss]elf[- ]?[iI]mmolat [Cc]utt

[Hh]ead[- ]?[Bb]ang [Oo]verdos [Hh]opelessness [Pp]owerlessness [Hh]elplessness [Nn]egative[- ]?[Aa]ttitude

[Ee]motional[- ]?[Nn]egativism [Pp]essimism

[Dd]epress [Pp]assivity

[Ss]ad[- ]?[Aa]ffect [Ss]adness

[Dd]ecreased[- ]?[Aa]ffect [Cc]ognitive[- ]?[Rr]igidity [Ss]uicidality

[Ss]uicide[- ]?[Ii]deation

[Mm]ental[ -]?[Hh]ealth

[Mm]ental[ -]?[Hh]ealth[ -]?([Cc]ris[ei]s|emergenc) ([pP]sychiatric|[Pp]sychotic|[Ss]chizophren\w*|[Bb]ipolar|[Mm]ental\w* ([Ii]ll\w*|[Dd]isorder))[ -

]?([Cc]ris[ie]s|[Ee]mergenc|[Aa]cute) ([Cc]ris[ie]s|[Ee]mergenc\w*|[Aa]cute)[ -

]?([pP]sychiatric|[Pp]sychotic|[Ss]chizophren\w*|[Bb]ipolar|[Mm]ental\w* ([Ii]ll\w*|[Dd]isorder))

